# Challenges in defining Long COVID: Striking differences across literature, Electronic Health Records, and patient-reported information

**DOI:** 10.1101/2021.03.20.21253896

**Authors:** Halie M. Rando, Tellen D. Bennett, James Brian Byrd, Carolyn Bramante, Tiffany J. Callahan, Christopher G. Chute, Hannah E. Davis, Rachel Deer, Joel Gagnier, Farrukh M Koraishy, Feifan Liu, Julie A. McMurry, Richard A. Moffitt, Emily R. Pfaff, Justin T. Reese, Rose Relevo, Peter N. Robinson, Joel H. Saltz, Anthony Solomonides, Anupam Sule, Umit Topaloglu, Melissa A. Haendel

**Author notes:** **Contact author**: Melissa A. Haendel, Center for Health AI, University of Colorado Anschutz Medical Campus, Aurora, CO, USA.

## Abstract

Since late 2019, the novel coronavirus SARS-CoV-2 has introduced a wide array of health challenges globally. In addition to a complex acute presentation that can affect multiple organ systems, increasing evidence points to long-term sequelae being common and impactful. The worldwide scientific community is forging ahead to characterize a wide range of outcomes associated with SARS-CoV-2 infection; however the underlying assumptions in these studies have varied so widely that the resulting data are difficult to compareFormal definitions are needed in order to design robust and consistent studies of Long COVID that consistently capture variation in long-term outcomes. Even the condition itself goes by three terms, most widely “Long COVID”, but also “COVID-19 syndrome (PACS)” or, “post-acute sequelae of SARS-CoV-2 infection (PASC)”. In the present study, we investigate the definitions used in the literature published to date and compare them against data available from electronic health records and patient-reported information collected via surveys. Long COVID holds the potential to produce a second public health crisis on the heels of the pandemic itself. Proactive efforts to identify the characteristics of this heterogeneous condition are imperative for a rigorous scientific effort to investigate and mitigate this threat.

## Introduction

SARS-CoV-2 emerged in late 2019 as the third human coronavirus identified in the 21st century. As of early 2021, new impacts of the virus are still being identified. The virus initially targets epithelial cells, endothelial cells, alveolar macrophages (via ACE2 proteins and the TMPRSS2 protease) causing symptoms attributable to the lungs, digestive tract, kidneys, heart, brain, and other organs.^1,2^ Additional research has begun to explore viral presence in other tissues that exhibit ACE2 and TMPRSS2 expression; these include skeletal muscle, smooth muscle, bone, cartilage and synovia.^3–6^ Collectively, these symptoms constitute coronavirus disease 2019 (COVID-19). Individual symptoms and disease severity vary widely among patients, with some patients developing mild or even asymptomatic infections, while others experience acute respiratory distress syndrome (ARDS), sepsis, and other life-threatening conditions.^7,8^

As more information about patient recovery has been collected, and as pathophysiologic mechanisms are revealed, a wide range of outcomes following acute COVID-19 have emerged. Some patients experience residual symptoms and others develop new symptoms long after the initial infection. These symptoms can present across a wide range of organ systems and tissues. Given the timeline of SARS-CoV-2’s emergence, studies to date have tracked patients’ clinical course up to six months post-infection,^9–14^ but anecdotal reports are available describing patients with ongoing symptoms as long as a year post-infection.^15,16^ Symptoms experienced after the acute illness represent a significant challenge for patients, physicians, and society as a whole. The causes, patient profile, and even symptom patterns associated with Long COVID remain difficult to isolate, and the natural history of this condition remains uncharacterized.

### Post-Acute Sequelae after Other Infections

The fact that some COVID-19 patients experience symptoms following recovery from acute infection is not unexpected. Other infectious diseases, including Epstein-Barr Virus,^17^ Giardia lamblia, Coxiella burnetii, Borrelia burgdorferi (Lyme disease) and Ross River virus,^18^ are also associated with an increased risk for post-infectious sequelae. These sequelae include symptoms such as disabling fatigue, musculoskeletal pain, neurocognitive difficulties, and mood disturbance.^17–19^ Chronic fatigue syndrome (CFS) is frequently preceded by a viral infection.^20^ However, although these sequelae are well documented, they are still not well understood, and the molecular mechanisms underlying these post-acute presentations have yet to be elucidated.

Post-infectious sequelae have also been documented following infection by other coronaviruses. A subset of patients with severe acute respiratory syndrome (SARS), caused by the coronavirus SARS-CoV, and Middle-Eastern Respiratory Syndrome (MERS), caused by the coronavirus MERS-CoV, were observed to experience persistent or new-onset symptoms, including fatigue,^21^ following recovery from the acute infection.^21–23^ For SARS, follow-ups have been conducted up to 15 years post-infection. In addition to fatigue, studies reported effects on lung health and capacity,^24–27^ psychological health,^21^ bone health,^27^ and lipid metabolism,^28^ with the latter two attributed to treatments involving large doses of steroids.^27,28^ Most of the improvements among SARS patients occurred within the first one to two years following infection,^27,29,30^ although some patients continued to experience decreased quality of life for more than a decade following the acute illness.^28^ Though follow-up studies in MERS patients are more sparse, effects on pulmonary function were observed at one year post-infection, with patients who experienced more severe cases at greater risk for long-term effects.^31^

### Post-Acute Sequelae Following COVID-19

While post-acute sequelae are not an unexpected outcome of SARS-CoV-2 infection, the number of people affected and range of symptoms associated with Long COVID is unprecedented. The multisystem nature of Long COVID compared to previously studied post-acute sequelae of human coronaviruses has raised questions about how to most effectively identify indicators of Long COVID. An analysis of 32 symptoms in patients with and without SARS-CoV-2 infection identified several symptoms that were enriched in patients with COVID-19 in comparison to other illnesses of comparable severity.^32^ After 30 days, loss of smell, loss of taste, memory loss, chest pain, and muscle weakness were the symptoms enriched in patients who were positive for SARS-CoV-2 at the time of their acute illness. The association between these symptoms and COVID-19 diagnosis fluctuated slightly at 60 and 90 days, with muscle weakness no longer associated at 60 days, difficulty concentrating emerging at 60 days, and confusion and bone or joint pain emerging at 90 days. Many of the symptoms most strongly associated with Long COVID are therefore distinct from those observed in post-acute SARS or MERS, and therefore may be challenging to identify based on research on other post-infectious sequelae.

Furthermore, regardless of whether they are unique to Long COVID, symptoms frequently reported by Long COVID patients are not assessed consistently across studies. A systematic review available as a preprint^33^ evaluated all research on Long COVID released prior to January 1, 2021, that included at least 100 patients; based on the 15 studies that met the inclusion criteria, the authors identified 55 symptoms of Long COVID. None of the most common symptoms were assessed by all 15 studies. They reported that the five most common symptoms evaluated in the literature were fatigue, headache, attention disorder, hair loss, and dyspnea. They also reported the frequency at which clinical measurements such as chest X-ray and biomarkers such as C-reactive protein and D-dimer were evaluated. The authors concluded that the symptoms of Long COVID are extremely heterogeneous, and that the assessment of these symptoms varies widely among studies.

However, Long COVID’s emergence has followed a different trajectory than that of most medical syndromes. Rather than building from a clinically determined framework of the illness, to date, much of the growing awareness of Long COVID and its symptoms has been driven by patient-led efforts.^34,35^ Observing residual or new symptoms months after experiencing COVID-19, patients have established online communities to provide support and identify similarities in their experiences.^36^ Some Long COVID patients who are also researchers have led efforts to systematically categorize the range of experiences associated with Long COVID. An extensive patient-led survey (Patient-Led Research Collaborative) performed deep longitudinal characterization of the Long COVID symptoms and trajectories in suspected and confirmed COVID-19 patients who reported illness lasting more than 28 days.^13^ Evaluating data from 3,762 respondents to 257 survey questions, this analysis documented 205 phenotypic features associated with Long COVID. The symptoms most frequently reported after 6 months were fatigue, post-exertional malaise, and cognitive dysfunction. Patients who reported symptoms lasting for longer than six months following acute infection experienced an average of 14 symptoms in month 7, and 86% of patients experienced relapses during the period assessed,with exercise, physical or mental activity, and stress reported as common triggers. The diversity of the symptoms reported by Long COVID patients underscores the urgent need to understand the natural history of COVID-19 following the initial infection in order to manage medical care of affected individuals.

Given Long COVID’s very recent emergence, no standard framework has yet been established for identifying and assessing associated symptoms or other clinical indicators. Most of the studies analyzed in the systematic review^33^ utilized a survey-based approach, meaning that they were able to analyze only symptoms identified *a priori* as concerns. These studies also varied in whether they included formerly hospitalized patients exclusively or a mixture of patients with mild, moderate, and severe cases of acute COVID-19. Long COVID can occur following either severe or relatively mild acute illness,^37^ and it has been suggested that the severity of acute illness affects the clinical course of Long COVID,^32^ as it does in SARS and MERS.^38^ Additionally, patients who are treated in the intensive care unit (ICU) would be assumed to be particularly likely to experience ongoing health challenges due to the well-documented occurrence of post-intensive care syndrome (PICS).^39^ Several different frameworks have been proposed to describe Long COVID cases, without any clear criteria emerging about how to define the condition or how to stratify patients. This ambiguity presents a concern as more and more data is collected: as of the end of 2020, at least 239 papers and preprints about the post-acute effects of COVID-19 had been released, and approximately 20 additional papers become available each month.^40^ These papers do not conform to a single definition of Long COVID and do not evaluate consistent symptoms or markers of the disorder (or constituent disorders). In addition, the differences between the common symptoms as identified in a systematic review of the literature^33^ compared to the patient-led assessment^13^ indicates that current research on Long COVID may fail to address the full diversity of and even the most significant symptoms identified by patients with lived experience of Long COVID. Additionally, because proactive self-report is an important component of the patient-led research collaborative, symptoms and experiences of persons with low access to and uptake of technology may be under-represented in Long COVID studies thus far.

In order to develop clinical management strategies to prevent or mitigate Long COVID, it will be essential for studies to use a unified definition of Long COVID and its subforms so that data from different studies can be integrated to provide the foundation for robust statistical inferences about risk factors for the development of Long COVID, as well as the natural history and response to treatments. Additionally, it is essential that survey-based research efforts to investigate Long COVID operate from a framework that addresses the symptoms most common among and most debilitating to Long COVID patients. A rigorous framework for evaluating Long COVID will also help to elucidate the organ systems involved in the disease and its sub-forms; such a framework could help to distinguish for example pulmonary versus cardiovascular syndromes and whether these are interrelated.. In this analysis, we present methodologies, findings, and perspectives related to the extraction of data from the literature, from an extensive patient survey, and from the NCATS N3C Data Enclave (covid.cd2h.org/enclave) to provide guidance towards defining and identifying symptoms and patient variables that must be considered while designing and developing studies of Long COVID. Given that Long COVID is poised to produce an additional public health crisis on top of the COVID-19 pandemic,^36^ rapid harmonization of existing data and the integration of this information into new efforts to characterize Long COVID will be the critical next steps in responding to this looming threat.

## Methods

### Literature Review

In order to explore how Long COVID is currently being characterized and reported, we conducted an exploratory landscaping review of the literature. The results of this search will inform a future, more systematic review of this topic. In addition to searching PubMed (MEDLINE), we included searches of specialized databases (e.g.,CoronaCentral; WHO Global Literature on Coronavirus Disease) and relied on expert recommended key articles, with snowball techniques to find similar studies. Both published articles and preprints were included for abstraction. The questions we explored in the review were: for observational studies of Long COVID, how are studies characterizing Long COVID, and what outcomes are reported and/or associated with this syndrome? In addition we explored whether any COVID-specific measures or tests have been developed or validated, whether any patient subgroups or medical specialties report unique signs or symptoms, and what patient-reported and patient-centered outcomes were reported **(Supplemental Table 1)**. While specific Inclusion and Exclusion criteria were not developed, we did exclude papers discussing only rehabilitation therapy, mortality or hospitalization, as these were not outcomes specific enough to Long COVID.

**Table 1.** General definitions of Long COVID used in the literature. The 39 papers and preprints reviewed could be binned into four general categories in their operationalizations of Long COVID.

| **Characterization of Long COVID** | **# of studies reporting** | **References** |
| --- | --- | --- |
| COVID-19 Clinical Course (& Related) | 16 | 14,32,46,47,57,59,73–82 |
| COVID-19 Recovered Patients (& Related) | 8 | 56,58,83–88 |
| Long COVID | 10 | 13,60–68 |
| Post-Acute Covid-19 Syndrome, | 5 | 52,69–72 |

The newly emergent nature of Long COVID and lack of definition complicate traditional search methods. Each study abstracted was analyzed to identify the relationship of participant recruitment in the study to formal definitions of Long COVID that have been proposed. This analysis required evaluating how the duration of long-term symptoms was defined relative to the acute illness and whether patients were selected or stratified based on variables related to clinical course. Due to the proposed definitions at the time of analysis, the variables considered were: definition of onset of disease course (e.g., diagnosis, positive test, hospitalization), time elapsed since onset (as defined in each manuscript), patient-reported symptoms or clinical measures assessed, and tests or measurements reported or developed.

### Formal Definitions Used for Comparison

Long COVID can be broadly defined as delayed recovery from an episode of COVID-19 and is characterized by lasting effects of the infection, e.g., persistence of symptoms or onset of new chronic diseases, for far longer than would be expected.^41^ Although no firm criteria have been established to define the post-acute period or sub-categories within Long COVID, several sets of guidelines have been proposed for the classification of COVID-19-related disease phenotypes, and these criteria were compared to the definitions used in the literature. For example, a recently proposed public health framework classifies SARS-CoV-2-related disease into three categories.^42^ The first is acute COVID-19, or the disease most commonly associated with acute SARS-CoV-2 infection. The second category includes Multisystem Inflammatory Syndrome in Children (MIS-C) and in adults (MIS-A), a less common presentation of SARS-CoV-2 infection characterized by hyperinflammation that can appear 4-6 weeks after viral infection.^43^ The third category describes late sequelae.^42^ In terms of defining study cohorts, adherence with this definition would therefore require a clinical diagnosis, rather than a SARS-CoV-2 test alone, in order to distinguish MIS-C/A and COVID-19.

Other frameworks break down the “late sequela” category into subtypes depending on either timing or disease natural history. For example, the United Kingdom’s National Institute for Health and Care Excellence’s guideline on long COVID provides two definitions of postacute COVID-19: (1) ongoing symptomatic COVID-19 for people who still have symptoms between 4 and 12 weeks after the start of acute symptoms; and (2) post-COVID-19 syndrome for people who still have symptoms for more than 12 weeks after the start of acute symptoms.^44^ Similarly, PACS has been defined operationally as extending beyond three weeks from the onset of first symptoms, and the term chronic COVID-19 has been proposed to refer to PACS cases where symptoms extend beyond 12 weeks;^37^ these PACS definitions are consistent with the virological data available thus far.^45^

However, other criteria recommend defining the post-acute period as starting once a patient is discharged from inpatient acute care for those hospitalized longer than three weeks.^45^ Some authors go further and subdivide Long COVID into three groups:

1.patients who have experienced severe COVID with ARDS and experience long-term respiratory symptoms dominated by breathlessness;

2.individuals with milder initial disease who were not necessarily hospitalized during the acute infection but but who present with a multisystem disease with cardiac, respiratory, or neurological manifestations of end-organ damage; and

3.people who have persistent fatigue and other symptoms but with no evidence of organ damage.^46^

### Terminological Extraction from the Literature

We reviewed patient-reported symptoms reported in the literature (or caregiver-reported symptoms in the case of one pediatric study^47^) and created a table row for each symptom in each publication. We then used a Python script to extract symptoms;. symptoms remained exactly as described in the manuscripts except to adjust for capitalization, punctuation, plurals (e.g., headache versus headaches), spelling in British versus American English (e.g., dyspnoea versus dyspnea), and to standardize labels assigned to specific measures. The identifiers for specific assessments used were as follows: FLU-PRO for InFLUenza Patient-Reported Outcome,^48^ EQ-5D-5L for the 5-level EQ-5D,^49^ EQ VAS for the EQ visual analogue scale,^49^ and mMRC Dyspnea Scale Scores for Modified Medical Research Council Dyspnea Scale Scores.^50^ We tabulated the relationships between publications and the symptoms they reported; we then manually mapped symptoms to one or more body systems and visualized the result using a Sankey diagram (**Figure 1**).

**Figure 1.**
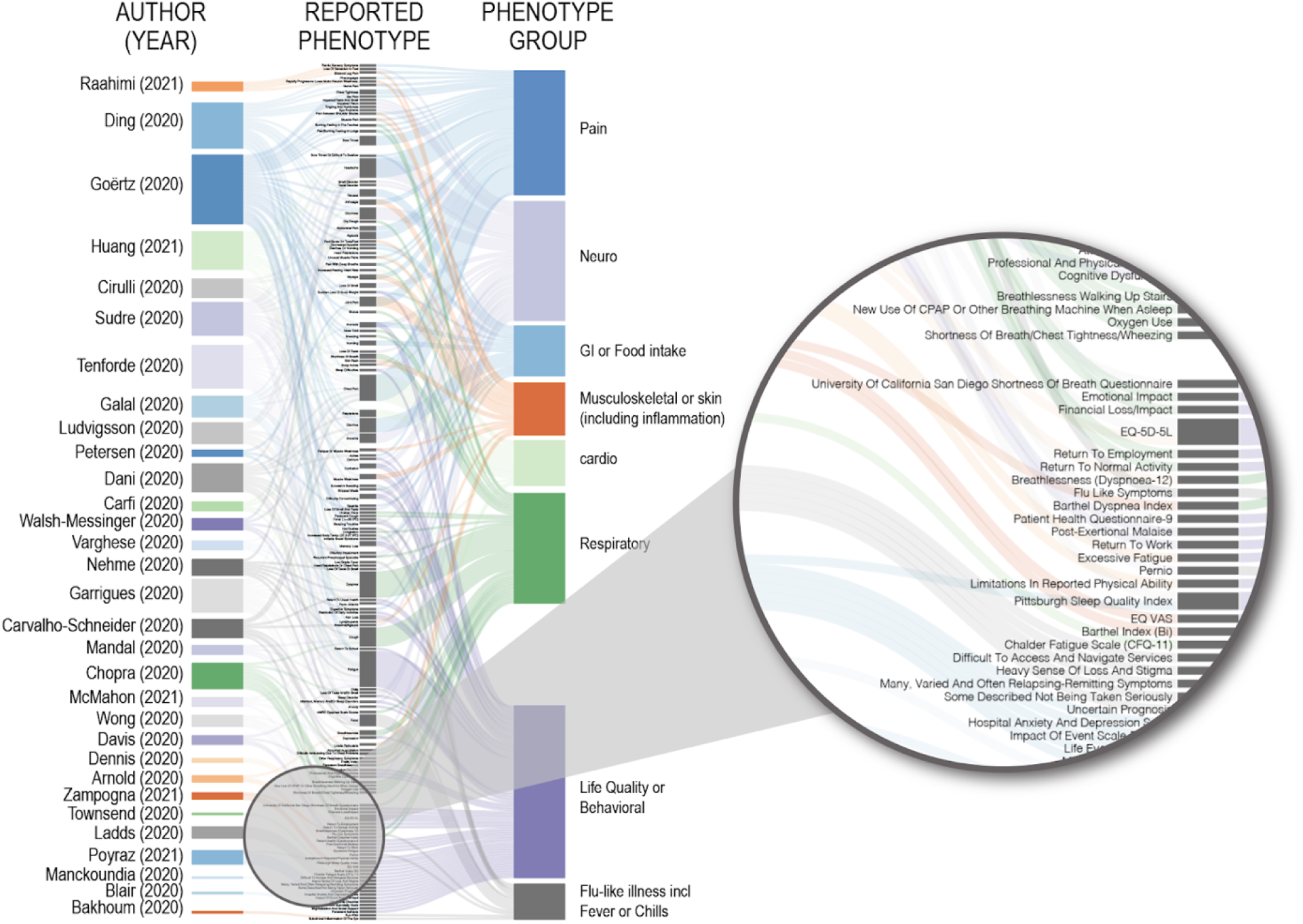
Heterogeneity of reported phenotypes for post-acute COVID-19 sequelae. Clinical and patient-reported symptoms, time course, and patients counts were extracted from the literature (see Supplemental Table 1. The author and year associated with each publication is provided in the first column. The second column indicates the exact phenotypes reported in each study, corresponding to symptoms and clinical indices. Symptoms and indices are categorized into phenotype groups. Most of the 142 symptoms or indices reported were unique to a single study. Examples of terms used are magnified in the pull-out. Supplemental Table 1 contains the literature extracted.

**Figure 2.**
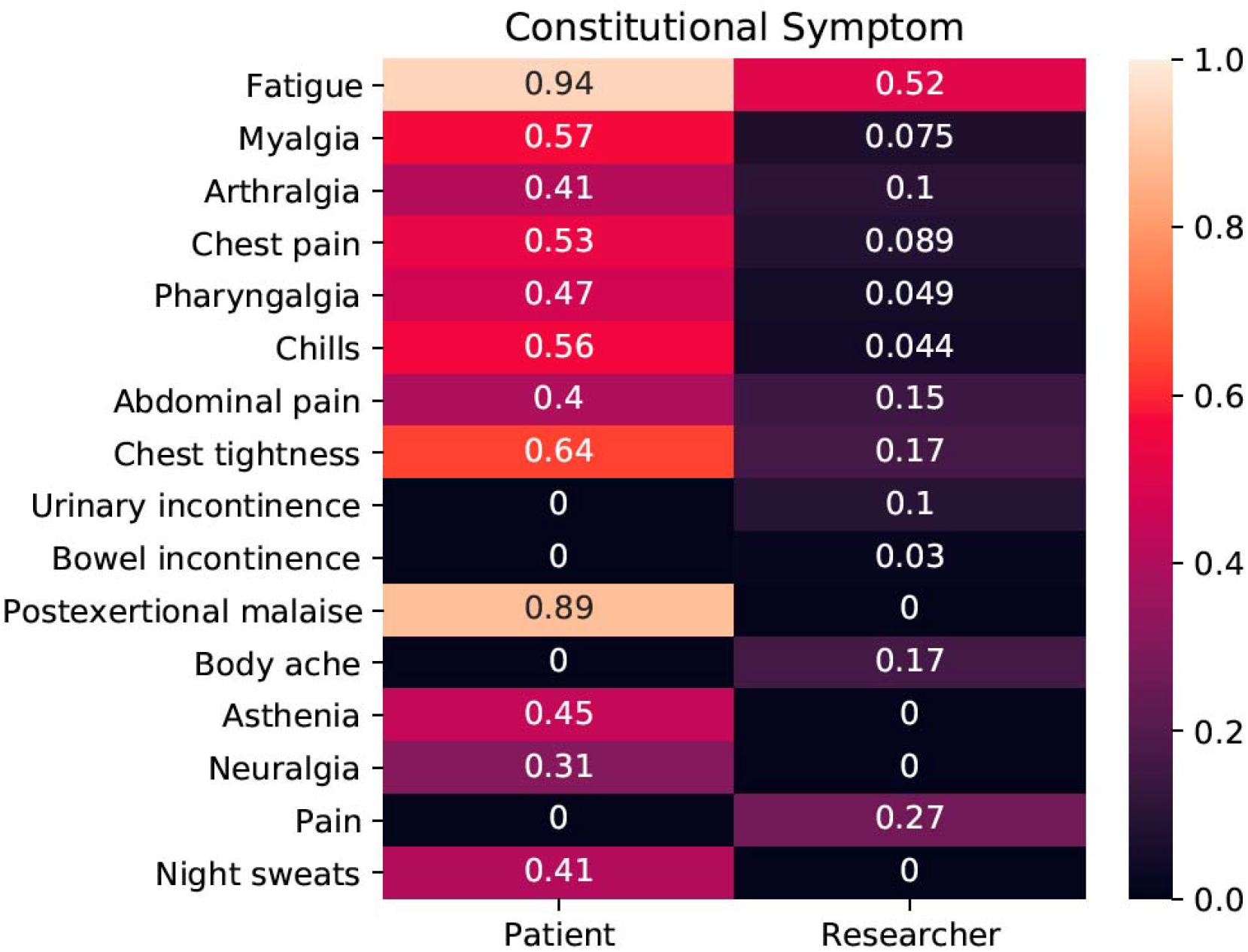
Average frequency of constitutional symptoms (specific terms descending from HP:0025142, Constitutional symptom, which is defined as a symptom or manifestation indicating a systemic or general effect of a disease and that may affect the general well-being or status of an individual). Frequencies are given separately for the 19 researcher-led studies and two patient-led studies.

### Ontological Coding of Literature and a Patient Survey

The Human Phenotype Ontology (HPO) provides a standardized vocabulary of over 15,000 terms to describe phenotypic abnormalities observed in human disease.^51^ In our review of the literature, we identified studies that also contained a description of the counts of affected individuals who displayed specific phenotypic features. We manually curated the mappings between literature-reported signs and symptoms and HPO terms. Overall, 141 unique symptoms were identified of which 80 terms were curated from the originally extracted literature terms, and 112 terms were captured from the both of the patient-led survey questions/answers.^13,52^ These are available in **Supplemental Table 2**.

### Cohort Selection

We performed analysis of electronic health record (EHR) data in the N3C Secure Data Enclave (covid.cd2h.org/enclave) with the intention of identifying unique healthcare utilization patterns among COVID-positive patients that may differentiate them as Long COVID patients. To achieve this, we looked for patterns found only in COVID-positive patients compared to COVID-negative controls. Some patients were expected to be COVID-positive but non-Long COVID, so this analysis was expected to distinguish at least three categories: COVID-positive and Long COVID, COVID-positive and non-Long COVID, and COVID-negative.

We define COVID-positive as any non-deceased patient in the N3C enclave with an ICD-10-CM diagnosis code for COVID (U07.1) or a positive PCR, antibody, or antigen test for COVID (*n =* 905,592). We define COVID-negative as any non-deceased patient in the N3C enclave with at least one negative PCR, antibody, or antigen test for COVID who is not also in the positive group (*n =* 2,473,206). We then further narrow the set of patients whose data is used for analysis in the following ways:

- We require all patients to have at least one year of history with their contributing health care system.
- For COVID-positive patients, we require that at least 90 days have passed since their COVID index date (minimum date of diagnosis or positive test).

Applying these restrictions resulted in a case (positive) cohort of 314,237 patients, and a control (negative) cohort of 1,917,935 patients.

We then employed the R package MatchIt^53^ to perform nearest-neighbor propensity matching on the positive and negative patients, at a ratio of 2:1 (control:case). The following factors were used in matching: age, sex, race, site (exact match required), and comorbid conditions (diabetes, chronic kidney disease, congestive heart failure, peripheral vascular disease, chronic pulmonary conditions). A patient was defined as having a comorbid condition if they had two or more ICD-10-CM codes equating to that condition in their EHR data. Two sites (representing 14,222 cases) were removed from the matching process due to a significant amount of missing data required for matching. Additionally, 2,311 cases were dropped because they were not able to be matched with two controls at their same site. This resulted in a final case set of 297,704 patients, and a final control set of 595,408 patients. The case set was further split into two groups: cases who were hospitalized for COVID (*n =* 51,903) and patients not hospitalized for COVID (*n =* 245,801).

We opted to model COVID-related healthcare utilization patterns among the cases and controls by counting occurrences of COVID-and Long COVID-related diagnoses (See “Long COVID Concept Sets,” below) for each patient before and after their COVID index date. (Controls were assigned their matched case’s index date.) Diagnosis occurrences were counted across an equal time period before and after the patient’s COVID index, based on how many days have passed since the COVID index. We ignored diagnoses occurring in a “buffer” period of 60 days before and after the COVID diagnosis, to attempt to differentiate “post-COVID” from active COVID.

After representing the data as a matrix of pre-and post-diagnosis conditions, we applied nonnegative matrix factorization in order to extract conserved co-occurring sets of diagnoses that best represent the cohort. The result of this step is a data-driven representation of which sets of diagnoses occur together. We then compared the change in frequency of these diagnoses before and after COVID to identify potential signatures of Long COVID.

### Long COVID Concept Sets

Concept sets were obtained by mapping a subset of the manually curated HPO concepts to OMOP concept identifiers within the Conditions domain (**Supplemental Table 3**). These mappings were obtained using OMOP2OBO.^54^ OMOP2OBO is an algorithmic framework designed to generate clinically meaningful mappings between Open Biomedical Ontologies (OBO) and standard clinical terminologies in the OMOP common data model. Using version 1.0.0 of the mappings, each of the HPO concepts were processed and all reasonable matches returned. HPO concepts unable to be mapped using OMOP2OBO were manually mapped using Version 1.12.0.6.210309.1608 of the Athena -OHDSI Vocabulary Repository,^55^ which at the time of mapping was populated with OMOP Vocabulary version: v5.0 26-FEB-21. All manual mappings were discussed with one or more professional ontologist and/or clinical phenotyping experts. Upon ingestion into the N3C Enclave, HPO concept sets were extended to include all descendant concepts for each included OMOP concept identifier. Each of the completed concept sets received an additional round of review by clinical domain experts within the Enclave prior to use in classifying the cohorts as described above.

**Table 3.**
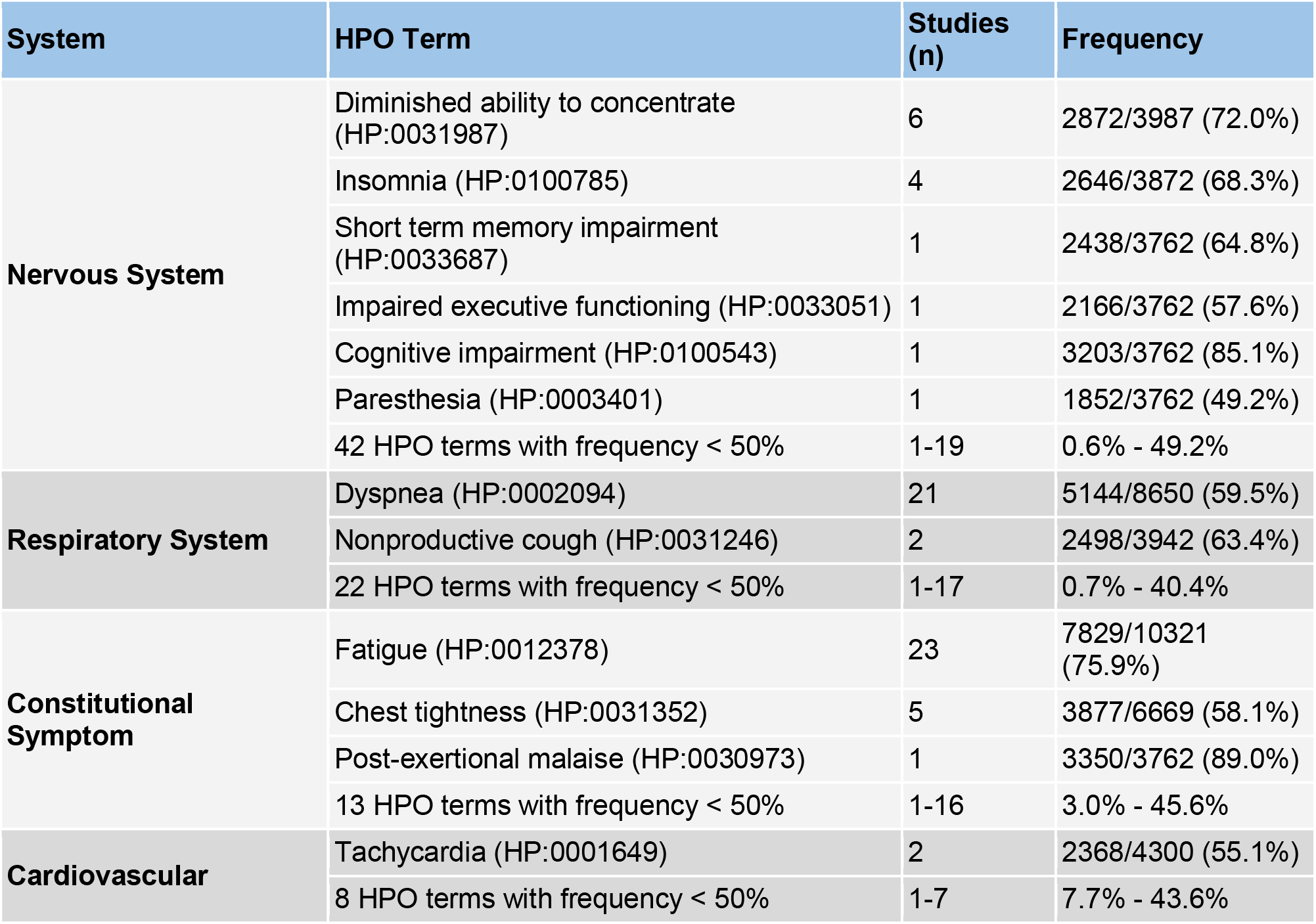
Summary of selected phenotypic manifestations in 21 studies (including two patient surveys) in post-acute COVID-19.

## Results

### Literature Review

The analysis of 39 studies revealed a variety of criteria were used to identify and evaluate patients with post-acute COVID-19 sequelae. With nearly as many definitions as studies, it is clear that there is no agreement on the definition of Long COVID (**Figure 1**). Studies differed in how they referred to the phenomenon studied. Some referred to it as Long COVID or using a similar term such as post-acute COVID-19 syndrome, whereas others discussed the clinical course or patient recovery without mentioning Long COVID specifically. These definitions fell roughly into four categories (**Table 1**). Most studies refer to their patient recruitment in terms of recovery (e.g., “COVID-19 survivors”^56^ or “discharged COVID-19 patients”^57^) or clinical course (e.g., “medium-and long-term consequences”^58^ or “delayed return to usual health”^59^). A number of studies did refer to their participant groups using terms like “Long COVID”,^13,60–68^ “post-acute COVID-19”,^69,70^ “post-COVID syndrome”,^71^ or “post-acute COVID-19 syndrome”,^72^ but these terms were not standardized among studies. A few studies^46,60^ acknowledged the proposed distinction at 12 weeks post-infection between post-acute COVID-19 and chronic COVID-19,^37^ but otherwise the definitions used typically did not refer to any proposed operationalizations of Long COVID. Therefore, while operational definitions of the constituent components of Long COVID have been proposed,^37,42,44^ reviewing the Long COVID literature revealed that they are rarely used when describing cases or identifying study cohorts.

Moreover, the existing operational definitions of Long COVID differ in important ways, many of which are not differentiated by existing studies. For example, one framework^46^ subdivides Long COVID patients into three groups based on whether their long-term symptoms are primarily respiratory in nature following severe COVID-19 with ARDS, whether they present with a multisystem disease with cardiac, respiratory, and/or neurological manifestations of end-organ damage, or whether their primary symptoms are persistent fatigue and other symptoms that do not necessarily indicate organ damage.^46,89^ In the literature analyzed, this definition was never used to define cohorts. Many studies included patients with acute infections that varied in their severity, including both inpatient and outpatient convalescents (e.g., ^82,85^). Additionally, of the studies available thus far, data directly assessing organ damage is rarely collected, and the concept of organ damage itself has not been operationalized in this context.

Other efforts to define Long COVID identify the severity of the acute phase as an important consideration in determining the onset of the post-acute phase. Specifically, for individuals hospitalized for more than three weeks following symptom onset, some definitions identify the post-acute period as starting once the patient is discharged from inpatient acute care.^45^ In the literature surveilled, most studies recruited and assayed patients based on time elapsed from a COVID-19-related milestone, but what the milestone was varied widely. Some studies use the date of diagnosis or positive test, others the onset of symptoms, others hospital discharge, and others by even broader criteria (e.g., patients with suspected or confirmed COVID-19 in the past). Many studies used a relatively precise window for patient assessment (e.g., 30 to 45 days after diagnosis^65^ or 14 to 21 days after symptom onset^59^), while others included participants at various distances from acute SARS-CoV-2 infection under the umbrella term of Long COVID.^46^ In the latter case, these patients could fall under either the Long COVID (PACS) or chronic COVID-19 definitions if using a 12-week cutoff.^37,44^ Because the relationship between infection, symptoms, and viral clearance occupies a wide distribution,^62,90,91^ this heterogeneity among and sometimes within studies could introduce significant variability in disease course within and among patient cohorts.

Finally, studies varied wide in the terminology used to describe patient-reported symptoms. Comparing symptoms described across the literature reviewed revealed 142 unique terms related to symptoms, including scales used to assess symptom profiles (e.g., the University Of California San Diego Shortness Of Breath Questionnaire) or other dimensions of recovery (e.g., 5-level EuroQoL 5-Dimensions for quality of life) (**Figure 1**). The most commonly evaluated symptoms were fatigue (15 studies), dyspnea (11 studies), chest pain (11 studies), and headache (8 studies). In many cases, studies assessed similar symptoms but differed in the nomenclature used. For examples, the studies analyzed included a mixture of reports of ageusia,^32,52,79^ anosmia,^32,52,79,82^ anosmia/ageusia,^76^ loss of smell,^59,68^ loss of taste,^59^ loss of smell and taste,^66^ loss of smell or taste,^65^ and loss of smell and/or taste.^77^ While in many cases there are parallels among studies (e.g., studies reporting anosmia and loss of smell are likely to be asking the same or similar questions of patients), the lack of a strict definition prevents straight-forward symptom matching across analyses. Further, there seemed to be limited surveying of neurological and systemic symptoms in some cases, hence the absence of common symptoms like cognitive dysfunction or “brain fog", sensorimotor symptoms, and post-exertional malaise. This is where standard use of a full terminology such as HPO would be useful to create expressive and consistent meaning across studies.

Therefore, the literature indicates that at present there is little consistency among studies in definitions of Long COVID, including the symptoms analyzed. Few studies use terminology with a proposed, narrow-scope definition such as Greenhalgh and colleagues^37^ definitions of PACS and chronic COVID. Instead, studies typically define a period of time to investigate symptoms agnostic of how this factors into the broader conversation on the disease. The exception is studies that state they are investigating Long COVID, which use a wide variety of definitions. The same is true for post-acute COVID-19 or post-COVID syndrome, which are typically not explicitly tied to working definitions or explicit disease phenotypes. Among studies, patient inclusion criteria can be based on any number of relevant milestones from the acute phase, and only a subset of studies separate patients based on the severity of disease they experienced in the acute phase. Finally, no standardized terminology is used for patient-reported symptoms, and studies often report symptoms using similar but non-identical terminology. Thus, the literature analysis suggests significant heterogeneity among studies with respect to how they define cohorts of interest and analyze the experiences of patients experiencing Long COVID.

### Ontological Analysis of Literature

From the results of the above exploratory review, candidates were selected to comprise a cohort of studies for further abstraction and analysis. Because of the poor reporting by and heterogeneity within our initial set of literature, the numbers of studies in this cohort is much smaller than the set of studies summarized above. This highlights the need for improved quality and reporting of even small cohorts. Details such as the specific definition used to identify patients with COVID-19 (e.g., a PCR test versus a clinical diagnosis), hospitalization status (outpatient versus inpatient versus ICU), the severity of illness represented among patients in the cohort, and the number of patients presenting with each symptom or other clinical measure are important to efforts to compare results across studies.

Here, 21 studies, including 20 published studies^14,47,52,56,58,59,63,66,72,73,75–77,79,92–97^ and one preprint,^13^ were chosen for the in-depth phenotypic analysis using HPO. A total of 154 different phenotypic abnormalities could be encoded using HPO terms. **Table 3** provides an overview of the most commonly observed abnormalities in four major categories, and **Supplemental Table 2** contains information about all 154 terms. The studies investigated and reported the phenotypic features in a very heterogeneous fashion. Only one abnormality, dyspnea (shortness of breath), was reported in every study. 95 terms were reported on only a single study.

## EHR Analysis

### Transforming the HPO codesets

For the EHR analysis, we focused on 77 HPO annotations commonly used in the literature. Of these, 76 were successfully mapped to at least 1 OMOP concept identifier within the Condition domain (min=1, max=84, median=3). The unmapped HPO concept, increased circulating brain natriuretic peptide concentration (HP:0033534), could not be reasonably aligned to an OMOP concept identifier within the Condition domain. When expanding each OMOP concept identifier to include its descendant concepts, the total number of OMOP concepts used was 7,542 (4,694 unique) and the median number of OMOP concept identifiers mapped to each HPO codeset was 16. The largest HPO codeset sets were paresthesia (HP:0003401; n=1,606 concepts), pain (HP:0012531; n=1,399 concepts), skin rash (HP:0000988; n=505 concepts), and anxiety (HP:0000739; n=355), which was not unexpected given the variability in the clinical presentation (e.g., severity, duration, and location) of the conditions associated with these concepts.

### Defining EHR phenotypes, including/excluding HPO codesets

The 297,404 patients in the final case group represent the pool of patients from which we have the potential to detect Long COVID (**Figure 3**). 85,912 of these patients had at least one instance of the identified HPO codes in their post-COVID period (and thus may be more likely to have Long COVID). Slightly more than half of these patients showed an increase in HPO codes after their diagnosis, with the largest shifts observed in hospitalized patients. Reduced dimension representation of the data suggested that HPO groups related to pain, anxiety/depression, and respiratory ailment. Further analysis will be required to determine which clusters of HPO codes are potentially indicative of Long COVID, allowing us to further stratify patients.

**Figure 3.**
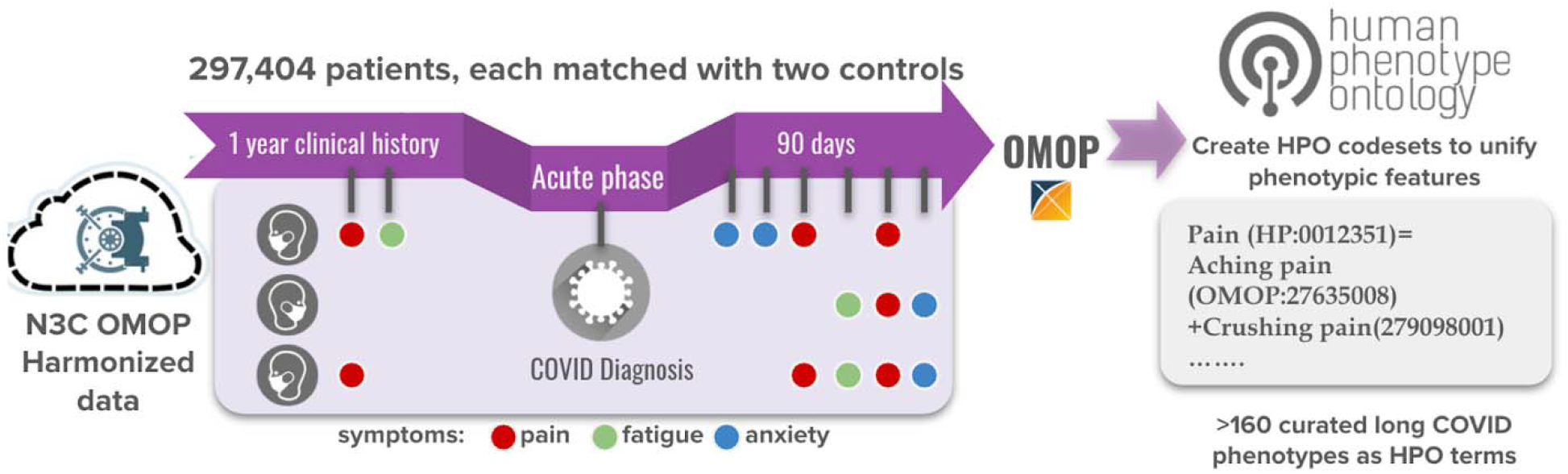
Schematic illustrating the method used to identify patients for Long COVID analysis, mapping of these patients’ data to HPO via OMOP2OBO codesets, and looking for patients with HPO phenotypic features from the mapped data to define a potential Long COVID cohort.

## Discussion

The analyses described above demonstrate the heterogeneity both in symptoms associated with Long COVID and in assessments and definitions used to study Long COVID present in the literature as well as an EHR-based approach for identifying natural language data associated with potential Long COVID patients available in N3C.

### Sources of Variance in Defining Long COVID

The literature review revealed a wide variety of terms used in describing patient cohorts used for studies of symptoms occurring after the acute phase of COVID-19. Most studies do not seek to assign their patients to a particular diagnosis or operational definition, although several referred to the definition from Greenhalgh et al. (2020)^37^, which is consistent with the virological data available thus far.^45^ There are a number of dimensions in which the existing literature varies in efforts to operationalize definitions of Long COVID. These differences are expected to vary in their effects. An important goal in the next phase of Long COVID research needs to be identifying the most critical considerations in defining patient cohorts.

#### Ambiguity in Defining the Acute Infectious Period

Long COVID is typically defined based on an elapsed acute infectious period, but at present, the relationship between the timing of COVID-19 symptoms relative to SARS-CoV-2 infection is not well understood.^98^ One early study examining viral load in hospitalized patients reported that viral shedding continued for at least 28 days following symptom onset in some patients.^99^ Another study reported that the median period between a patient’s first positive PCR test and cessation of viral shedding was 17 days and that up to 70% of patients were still symptomatic when their viral shedding ceased.^90^ However, viral shedding (e.g., the presence of detectable SARS-CoV-2 virus in samples such as nasopharyngeal swabs) does not necessarily indicate the presence of replication-competent viral particles. Viable viral particles have been detected from 6 days prior to up to 9 days after symptom onset.^100–102^ Patients have also been observed to test positive by PCR following a negative test,^103–105^ but the virus could not be cultured. Both asymptomatic and symptomatic patients with retest-positive COVID-19 have been identified.^103^ Even in individuals whose nasopharyngeal swabs produce negative PCR results, some test positive for SARS-CoV-2 in the intestine.^106^ These results therefore suggest that after the initial infection, patients shed non-infectious, degraded viral particles.^104^

In Long COVID, this relationship is further complicated by the fact that many patients who report symptoms of Long COVID lack a formal diagnosis. Due to the scarcity of tests in many places at the beginning of the COVID-19 pandemic, many patients who had suspected COVID-19 were never tested for the presence of the SARS-CoV-2 virus.^107^ In current studies, there is significant variability in the inclusion/exclusion criteria used for patient recruitment in terms of COVID-19 test status. While some studies require a positive test, others recruit patients with either a confirmed or suspected diagnosis (**Table 1**). Furthermore, some studies fail to specify whether the tests used for selecting patients are PCR-based, serum antibody based, or a mixture of the two. This distinction is important because the rate of false positives and false negatives is much higher in the antigen/antibody tests,^108–111^ meaning error rates may vary among studies. This limitation presents challenges for clinicians in determining the likelihood that patients with non-specific symptoms have Long COVID, and also presents difficulties for large-scale efforts to characterize symptoms associated with COVID-19 and Long COVID.^32,112^

#### Initial EHR characterization of a potential Long COVID Patient Cohort

There is not currently an ICD-10-CM diagnosis code for long-COVID; thus, our ability to find patients with long-COVID using structured EHR data is limited. Lacking an ICD-10-CM code, we utilized the HPO terms curated from the literature and patient surveys to refine the potential cohort based by looking for patients with at least one of these specific HPO terms. The patients characterized in **Table 4** represent a base population from which EHR analysis *may* be able to identify Long-COVID. These are patients who had COVID, have enough pre-COVID longitudinal data to enable us to compare their healthcare utilization pre-and post-COVID, and have had enough time pass since their COVID diagnosis to be out of the acute phase. While we cannot say with certainty that the patients who reported one or more long-COVID symptoms have long-COVID, as shown in **Table 4**, the characteristics of this group are significantly different from those cases lacking a reported symptom. This cohort would be an ideal group for deeper phenotyping, leveraging additional data sources such as features derived from free-text notes in the EHR, imaging, or claims data.

**Table 4.** Defining a cohort of potential Long-COVID patients. Comparing characteristics of non-deceased COVID patients with >= 1 year pre-COVID longitudinal data and >=90 days since COVID diagnosis (column 1); and COVID patients with >= 1 year pre-COVID longitudinal data, >=90 days since COVID diagnosis, and an instance of a Long COVID phenotypic feature >= 60 days after their COVID diagnosis (column 2).

| | | All qualifying cases | All qualifying cases with Long COVID HPO code $\geq 60$ days post-COVID | p |
| --- | --- | --- | --- | --- |
| n |  | 211,792 | 85,912 |  |
| Sex (%) | Female | 119,843 (56.6) | 55,478 (64.6) | p <0.001 |
|  | Male | 91,930 (43.4) | 30,425 (35.4) |  |
|  | Unknown | <20 (0.0) | <20 (0.0) |  |
| Age | mean (SD) | 44.62 (21.12) | 49.84 (19.77) | p <0.001 |
| Race | Asian | 5,401 (2.6) | 1,954 (2.3) | p <0.001 |
|  | Black | 35,241 (16.6) | 17,634 (20.5) |  |
|  | White | 135,563 (64.0) | 52,570 (61.2) |  |
|  | Other/Unknown | 35,587 (16.8) | 13,754 (16.0) |  |
| Ethnicity | Hispanic/Latino | 26,896 (12.7) | 11,192 (13.0) | p |
|  | Not Hispanic/Latino | 159,843 (75.5) | 63,829 (74.3) | <0.001 |
|  | Other/Unknown | 25,053 (11.8) | 10,891 (12.7) |  |
| **Comorbidities (Pre-COVID)** | Diabetes | 22,169 (10.5) | 16,270 (18.9) | p <0.001 |
|  | Kidney disease | 11,385 (5.4) | 9,308 (10.8) | p <0.001 |
|  | Heart failure | 6,482 (3.1) | 6,074 (7.1) | p <0.001 |
|  | Pulmonary disease | 12,971 (6.1) | 12,682 (14.8) | p <0.001 |

#### Related and Concurrent Disorders

One major issue arising from the challenges to determining whether a patient has recovered from COVID-19 is that post-acute symptoms can also arise from different etiologies. One potential source of ambiguity come from PICS, which describes new or worsening cognitive, psychological, and physical limitations experienced by patients following discharge from an intensive care setting.^39^ Some impairments have been observed to persist for years after discharge, including pulmonary effects that are exacerbated by intubation and can persist for five years or longer and decreased ability to conduct activities of daily living that can last for 1-2 years.^113^ Therefore, symptoms of PICS could potentially be conflated with symptoms of Long COVID in patients who were ventilated and/or treated for COVID-19 in the ICU. Another possible source of long-term symptoms is the treatments used during the acute illness. In SARS, some of the most common post-acute sequelae are thought to be caused by treatment with corticosteroids.^27,28^ Therefore, the care received during the acute phase of the illness holds the potential to influence the clinical course of recovery, and therefore should be considered in efforts to identify signifiers of Long COVID.

While COVID-19 is a complex and heterogeneous multisystem illness, patients infected with SARS-CoV-2 can also develop distinct illnesses. A multisystem inflammatory illness has been observed in children and in some adults following acute infection with SARS-CoV-2. This syndrome, called multisystem inflammatory syndrome in children (MIS-C) and in adults (MIS-A),^43^ is characterized by hyperinflammation and can begin subsequent to host clearance of active SARS-CoV-2 infection.^42^ This condition is rare, with estimates of two in every 100,000 children in a descriptive analysis of MIS-C cases in New York State.^114^ This report also identified a median of 21 days from when children experienced COVID-19 (or an illness likely to be COVID-19) and when they were admitted to the hospital for MIS-C and that they were hospitalized for a median of 6 days.^114^ MIS-A has been reported only very rarely, with only 30 known cases as of October 2020.^115^ The importance of distinguishing the natural history of MIS-C/A from that of COVID-19 has been highlighted in some efforts to operationalize definitions of Long COVID,^42^ but at present, MIS-C/A is not widely discussed in the Long COVID literature, even though it too manifests in the post-acute phase of infection.

Similarly, preliminary findings suggest that patients with SARS-CoV-2 infection are at risk for chronic illnesses associated with post-viral sequelae. One example is that some presentations of Long COVID bear a resemblance to CFS, another chronic condition that is often triggered by a viral infection.^20^ The broad relationship between these known sequelae of viral infections and the specific pathogenesis of SARS-CoV-2 remains to be identified, although some mechanisms have been proposed^116^. In terms of characterizing the long-term sequelae of SARS-CoV-2 infection, they may introduce additional ambiguity regarding the specific outcomes associated with this particular virus compared to viral infections more broadly.

#### Organ Damage

One definition of Long COVID^46^ specifically highlights the potential importance of distinguishing long-term symptoms arising from organ damage from those arising from other etiologies. Given that a large number of Long COVID patients suffer from fatigue, which is associated with other post-viral syndromes but for which there are limited treatment options,^20^ identifying whether and when Long COVID patients have sustained long-term organ damage may provide additional options for treatment and understanding of the disease. However, few studies of Long COVID to date have conducted analyses elucidating the presence or extent of organ damage. Many assessments to collect evidence of long-term organ damage are intensive, meaning that their feasibility may vary with the strain on hospitals during the course of the COVID-19 pandemic. However, preliminary investigations of a number of organ systems have identified organ damage in Long COVID patients. These findings are also important because they highlight the possibility of asymptomatic Long COVID patients, who could sustain organ damage due to the SARS-CoV-2 virus that does not immediately present with symptoms. Therefore, an improved understanding of organ damage as an outcome of acute COVID-19 or as a long-term sequelae of the SARS-CoV-2 virus may present new options for patients experiencing persistent symptoms or elucidate new information about how the SARS-CoV-2 virus interacts with a range of organ systems.

#### Post-AKI CKD, Diabetes, and Long COVID Syndrome

During acute SARS-CoV-2 infection, diffuse endothelial injury, leads to end organ perfusion abnormalities and microthrombi. This reduced perfusion contributes to acute kidney injury (AKI), and possibly to new-onset diabetes.^117–119^ AKI, especially moderate/severe AKI, is a risk factor for the development of chronic kidney disease (CKD).^120^ Apoptosis, maladaptive repair, and fibrosis have been postulated as mechanisms involved in the transition from AKI to CKD.^121^ The kidney is an organ of interest in Long COVID because acute SARS-CoV-2 infection is associated with kidney injury.^122,123^ SARS-CoV-2-associated microvascular injury may cause perfusion abnormalities within the pancreatic islets, skeletal muscle, heart and or brain. In the islet, for example, microcirculation is essential for both glucose sensing and insulin secretion; abnormal islet capillary architecture and fragmentation contributes to beta cell dysfunction in type 1 and type 2 diabetes.^*124*^ Diabetes is a known contributor to CKD. Both CKD and diabetes are major risk factors for cardiovascular disease (CVD)^125^ and long term disability, which may overlap with the complicated picture of PASC.^126^

An unpublished investigation and a complementary published analysis provide evidence highlighting the relevance of kidney damage to medium-to-long COVID-19 outcomes. A pilot investigation (unpublished) was conducted on a subgroup of 35 COVID-19 AKI survivors who were admitted at Stony Brook University Hospital, NY between March and June 2020 and subsequently followed in a “Post-AKI COVID clinic.” Patients were observed at a 6-month follow-up to have a high incidence of persistently reduced renal function after moderate/severe AKI in the setting of hospitalization with COVID-19. *De novo* or progressive CKD was noted in 25.7% & 74.3% of cases based on estimated glomerular filtration rate (eGFR) + serum creatinine (SCr) and only SCr measures, respectively. A second study in a Swedish cohort^127^ similarly investigated kidney dysfunction following acute illness. In a group of 60 ICU patients admitted for COVID-19 infection, they found that inpatient AKI severity was associated with higher CKD stages at 3-to 6-month follow up.^127^ They found no differences between patients with CKD progression compared to those without progression in terms of demographics, comorbid conditions, or ICU admission characteristics.^127^ Similarly, in the unpublished study, neither inpatient AKI recovery nor a history of CKD prior to admission were associated with worsening renal function at follow up. Both of these analyses are limited due to a small sample size. Ongoing study at Stony Brook’s Post-AKI COVID clinic will include additional patients and longer follow-up and therefore should provide a more accurate estimate of CKD risk. It is not yet known whether inadequate renal repair after severe injury or persistence of SARS-Cov-2 in the kidney drives post-AKI CKD in COVID-19.

While AKI is an established independent risk factor for CKD,^128^ this association has not yet been extensively explored in the setting of COVID-19, given that the virus has been circulating for just over a year at this time and studies so far have mostly reported the persistence of renal dysfunction (AKD) at time of hospital discharge.^129,130^ Persistent organ damage is now considered part of the Long COVID syndrome,^45,131^ and kidney disease should be considered part of this syndrome. While these two studies are among the first reporting this association, further multi-center studies with larger sample sizes and with pathology data are needed to further analyze the relationship between AKI and development/progression of CKD in COVID-19.

#### Neuroimaging in Analyses of Long COVID

A variety of neuroimaging findings have been reported in COVID-19 patient populations, and efforts to better understand pathophysiologic origins and neuroanatomical correlates are ongoing. A number of studies have been carried out to characterize COVID-19 neuroimaging findings and associated neuropsychiatric symptoms, e.g. ^132–135^. There have been a few focused imaging studies that attempt to dissect neuroimaging correlates associated with specific symptoms; for instance, olfactory bulb abnormalities were characterized in an MR imaging study of COVID-19 anosmic patients.^136^ One study comparing 35 Long COVID patients to 44 controls found significant hypometabolism in the brain, including the olfactory gyrus, right temporal lobe (including the hippocampus and amygdala), the bilateral pons/medulla brainstem, and the bilateral cerebellum; notably, the clusters of hypometabolism were correlated with patient symptoms, including hyposmia and anosmia, memory and cognitive impairment, pain, and insomnia.^137^ There have also been additional suggestions that brainstem dysfunction might be involved in a variety of COVID-19 clinical manifestations. For instance, Yong^138^ cites a number of autopsy studies to support this hypothesis.^139,140^

#### Autopsy as a Means to Diagnose Long COVID

Autopsy analysis is an important method to obtain insights into the pathology associated with COVID-19 and the presence of the SARS-CoV-2 virus in diseased tissues. Kidney tissue provides a good example of the importance of autopsy analysis. AKI is very common in patients hospitalized with COVID-19 and is a major risk factor for mortality.^141–143^ Kidney autopsies or biopsies in patients with Covid-19 related AKI do not generally reveal suggestions of direct viral cytotoxic effects such as nuclear, cytoplasmic inclusions or with extensive tissue necrosis and inflammation.^144^ Autopsy and kidney biopsy tissue studies have indicated that acute tubular injury is the most common pathologic finding Covid patients with AKI or proteinuria.^145^ Collapsing glomerulopathy and thrombotic microangiopathy are also been associated with Covid 19 AKI in autopsy and biopsy studies.^145^

Multi-organ and especially renal tropism of SARS-CoV-2 has been observed in autopsy studies on COVID-19 patients. Puelles et al^1^ reported the presence of viral load and also viral RNA and proteins in the kidney using *in situ* hybridization and indirect immunofluorescence with confocal microscopy. In an autopsy study of 26 patients with COVID-19, Su et al.^146^ found clusters of coronavirus-like particles in the kidney tissue on electron microscopy and also detected positive immunostaining with SARS-CoV nucleoprotein antibody associated with injury patterns on light microscopy.^146^ Autopsy studies have also shown that SARS-CoV-2 infects and replicates inside pancreatic beta cells, reducing insulin-sensing functions of those cells.^147^ This direct infection of the pancreatic beta cells is likely a lead cause of metabolic dysfunction and glycemia after SARS-CoV-2 infection. The N3C database offers a unique opportunity to study glycemia before and after SARS-CoV-2 infection, as well as how new-onset diabetes may contribute to PASC effects on quality of life for both adults and children.

In a review of brain autopsy studies, summarizing 24 studies with results from 149 individuals chronic inflammation or neural changes typically associated with viral infections were found to be largely absent.^148^ Interestingly, in one recent study, megakaryocytes were found in cortical capillaries in 33% of brain autopsy cases examined.^149^ The authors thereof a) noted that this observation was consistent with other observers who have noted megakaryocytes^150, 151^ and b) suggest that these large cells could cause ischemic alternation in a distinct pattern and might be associated with COVID-19 neurological impairment.

The timing of autopsy is likely to be important in efforts to detect whether SARS-CoV-2 remains in tissue. In another study examining 42 postmortem samples of patients who died with COVID-19, no presence of SARS-CoV-2 was noted in analysis with immunofluorescence, electron microscopy or *in situ* hybridization of the kidney tissue.^152^ This study raised concerns about the method and timing of post-mortem tissue collection and processing, since a significant degree of autolysis was noted in the kidney tissue in this study.^153^ In a recent study of immediate (≤3 hours) post-mortem renal biopsies of 16 patients with COVID-19 and 5 control patients with sepsis, investigators reported that the presence nCoV2019 N-Protein was detected in proximal and distal renal tubules in 9 of 16 cases, out of which 6 of the 9 were confirmed by *in situ* hybridization. This finding supported the presence of SARS-CoV-2 in the kidney.^154^ However, SARS-CoV-2 E and N1/N2 genes were detected by RT-PCR of the kidney total RNA in only one case, and classical viral inclusions were not detected via electron microscopy. Therefore, while autopsy can serve as an important tool in looking for the presence of SARS-CoV-2 in tissues and the associated pathology, the methodology used for autopsy is critical to providing accurate insights into disease patterns.

#### COVID-19 and Quality of Life

The circumstances surrounding infection with SARS-CoV-2 and the pandemic itself are likely to have a significant impact on patients’ health. In particular, psychosocial health, nutrition, and physical fitness may all be impacted by the broader societal response to SARS-CoV-2. Early in the pandemic, the WHO released recommendations to support psychosocial health in light of the pandemic (Mental health and psychosocial considerations during the COVID-19 outbreak). Since then, many reports have indicated concern about a rise of psychosocial distress internationally.^155–158^ While unique psychological stressors are likely to affect patients who experience COVID-19 and especially those with more severe cases, the impact of a broader societal decline in psychological health (including addiction/substance abuse disorder) may be difficult to identify in studies that evaluate only COVID-19 patients with and without Long COVID. Similarly, viral infections can exacerbate pain and other chronic conditions,^159^ but these effects are not specific to SARS-CoV-2 even though they may appear that way depending on study design. Similarly, the conditions of the pandemic have reduced access to healthy food choices in some places^160,161^ and reduced opportunities for exercise.^162^ Social distancing also presents unique challenges to patients with substance use disorders; as loneliness and stress can make people more inclined to substance use.^163,164^ As governmental and societal responses to SARS-CoV-2 evolve, it is possible that quality of life and psychosocial reports from Long COVID patients may shift along with those of the population more broadly.^165^

#### Research Response and Measurement Problems

Because of the pandemic, there has been an incredible surge of research and a call for the surveillance of COVID-19 patients.^166,167^ Thousands of clinical trials are being registered, initiated and, in many cases, completed on COVID-19 treatment and prevention in the USA and across the planet.^168^ While this response is impressive, there are risks to rapidly planning and performing expedited clinical trials.^169^ For example, recent reviews of registered protocols have revealed methodologic flaws and a wide array of outcomes measures, particularly patient-reported outcome measures (PROMs), being used,^169–171^ most of which have not been vetted for relevance to COVID-19 patients.^172^ Moreover, the lack of available terminological standards greatly impede the ability to compare studies.

There are two obstacles to the design of clinical research in this area. First, there has not yet been any rigorous large-scale effort to characterize the constellation (incidence and breadth) of outcomes most important to Long COVID patients. Without this characterization it is not possible to design inclusion criteria for responsible clinical studies. A second and related obstacle is there have not yet been efforts to define the sets of core domains and outcomes for patients in future clinical studies. Heretofore the lack of uniformity in outcome measurement across clinical research creates multiple problems: it undermines the validity of this research, shows a lack of relevance to the patient perspective, and limits our ability to compare findings between studies or to pool data for meta-analyses.^139,173^

In an effort to reduce heterogeneity in outcomes measured across clinical trials, and to improve the clinical monitoring of patients, the development of core domain sets (CDS) and core outcome sets (COS) in specific health conditions has been routinely recommended.^174–176^ Core outcomes are instruments (e.g., EURoQOL scale, PROMIS Emotional Distress -Depression scale) that measure particular core domains (e.g., quality of life, depression, pain), the latter of which are specific symptoms or broader symptom categories. A CDS is an agreed upon selection of symptoms or symptom domains (categories) that should be measured and reported in all clinical trials for a particular health condition. A COS is defined as an agreed minimum selection of outcomes that should be measured and reported in all clinical trials for a particular health condition.^176^ CDSs must be developed prior to the development of COSs of measurement instruments. Given that the scientific community has only recently started to examine Long COVID, a CDS is the first necessary step. A CDS would increase the reporting of patient important outcomes in Long COVID, reduce the risk of selective outcome reporting,^177^ and increase the feasibility of conducting meta-analyses on such topics in the future.^174,177^ In relation to value-based health care, core domain and outcome sets are key to performing research that inform quality indicators related directly to patient outcomes and are routinely being used by national health-care organizations in the USA and abroad^178–184^ and in particular can be used in the measurement of quality of care in the COVID era.^185^

Some work has been done to create various types of CDSs and COSs for clinical trials of acute COVID-19.^170,186–188^ While this work is important for the acute period of COVID-19, these efforts do not focus on the long-term outcomes associated with Long COVID. To date no work has been done to explore what is important to patients with Long COVID. Without a CDS informed by a large sample of patients that had COVID-19, clinicians and clinical trialists will lack an essential assessment tool to adequately measure patient specific and patient important outcomes and changes across time. A CDS would provide a critical means of comparing results across trials, which is extremely difficult in the current conditions where many different PROMs are being used in many different samples of patients.

These problems undermine the relevance and usefulness of this evidence for decision-making, and the research does not focus on what is most important to patients. Because evidence suggests long-term effects of COVID-19 on health-related quality of life, working to identify the domains and corresponding measures (e.g., Patient Reported Outcomes Measurement Information System [PROMIS] item banks) that are most relevant to COVID-19 patients following the acute infection is urgently needed given the rapid expansion of clinical research in this group. The incidence of those with Long COVID will climb, and soon much clinical care and research will be directed at this group, as evidenced by the increase in research in the area.

### Importance of Defining Long COVID

Available evidence suggests that Long COVID is a substantial public health problem with severe consequences for affected individuals and society at large. Patients commonly report being emotionally affected by health problems related to Long COVID. In the United States, patients have reported mild to severe financial impacts related to acute or chronic COVID-19,^77,189^ This concern is underscored by reports that Long COVID patients experience increased disability related to breathlessness and decreased quality of life.^190^ Understanding the needs of these patients will allow for the development of healthcare, rehabilitation, and other resources needed to support their recovery.^191,192^ However, identifying patient needs is contingent on developing a research infrastructure that accurately assesses the natural history of this illness.

Given the heterogeneity of clinical presentations of individuals with prolonged clinical manifestations following acute COVID-19, it is likely that clinical management should be tailored to individuals. However, the clinical management of Long COVID remains challenging because there are no evidence-based guidelines. Existing studies do not always provide comprehensive information about the clinical course, and often present aggregated results for individuals with differing clinical courses, such as for instance severe COVID requiring admission to an ICU and moderate COVID requiring hospitalization but not care in the ICU. Existing literature is contradictory with respect to the natural history of Long COVID. For instance, one study found that persistent fatigue is independent of severity of initial infection, but another found that 10 of 16 individuals (63%) with severe acute COVID-19 but only 26/65 (40%) individuals with moderate COVID had persistent fatigue.^73^ It should be noted that available studies have investigated COVID-19 patients who have come to medical attention, and much less data are available at the population level about the extent of late sequelae.^42^

Most studies to date use survey-based methods to ascertain patient-reported symptoms of Long COVID, although some studies are beginning to use imaging and other technologies to identify the physical signs of organ damage. Vital signs are a third category of indicators that are likely to prove valuable in efforts to investigate Long COVID. Vital signs have several attractive properties for the study of COVID-19. Data are often available from prior to the illness allowing for pre-post comparisons and are routinely collected in affected and unaffected individuals allowing for case-control comparisons. Moreover, analyses of discontinuities in a vital sign’s trajectory of time are possible. An ecosystem where associations between patient-reported symptoms, data available in EHR, and results of simple and/or complex clinical assessments with Long COVID have been evaluated and standardized will introduce a positive feedback cycle where clinicians are able to collect the data needed for the elucidation of Long COVID phenotype.

While heterogeneity in the presentation of Long COVID has been identified, the specific variables influencing outcomes remain to be characterized. The number of syndromes within Long COVID and the extent to which symptom profiles, frequency of occurrence, and duration are unique to these groups remains to be explored. At present, however, data is not collected in a way to allow for these subtle differences to be parsed. In order to develop clinical management strategies to prevent or mitigate Long COVID, it will be essential for studies to use a unified definition of Long COVID and its subforms so that data from different studies can be integrated to provide the foundation for robust statistical inferences about risk factors for the development of Long COVID, as well as the natural history and response to treatments.

Among the reasons for needing an unambiguous definition of Long COVID is the need to make clear contrasts and comparisons between affected and unaffected people. In addition, a clear definition is necessary to understand whether or to what extent defining phenotypic features of Long COVID were present prior to COVID-19 illness in patients affected by Long COVID or potentially serving as controls in studies. The identification of appropriate unaffected people and pre-illness time periods for comparisons is foundational to advancing the state of the art in Long COVID research. It is imperative that patient-reported symptoms be taken into account alongside deep clinical characterization and large scale observational data such as in the N3C. However, all three sources of data are subject to biases and all sources are needed to provide a more complete picture of Long COVID characterization for individuals and populations.

## Ethics and Regulation

The N3C data transfer to NCATS is performed under a Johns Hopkins University Reliance Protocol # IRB00249128 or individual site agreements with NIH.

Use of the N3C data for this study is authorized under the following IRB Protocol:

| Site | IRB name | Exempted vs. approved | Protocol number |
| --- | --- | --- | --- |
| University of North Carolina | University of North Carolina Chapel Hill Institutional Review Board | exempted | 21-0309 |
| Stony Brook University | Office of Research Compliance, Division of Human Subject Protections, Stony Brook University | exempted | IRB2021-00098 |

The N3C Data Enclave is approved under the authority of the NIH Institutional Review Board for Protocol 000082 associated with NIH iRIS reference number: 546652 entitled: “NCATS National COVID-19 Cohort Collaborative (N3C) Data Enclave Repository.” Further information can be found at ncats.nih.gov/n3c/resources.

## Data Availability

The N3C Data Enclave (covid.cd2h.org/enclave) houses fully reproducible, transparent, and broadly available limited and de-identified datasets (HIPAA definitions: https://www.hhs.gov/hipaa/for-professionals/privacy/specialtopics/de-identification/index.html). Data is accessible by investigators at institutions that have signed a Data Use Agreement with NIH who have taken human subjects and security training and attest to the N3C User Code of Conduct. Investigators wishing to access the limited dataset must also supply an institutional IRB protocol. All requests for data access are reviewed by the NIH Data Access Committee. A full description of the N3C Enclave governance has been published;193 information about how to apply for access is available on the NCATS website: https://ncats.nih.gov/n3c/about/applying-for-access. Reviewers and health authorities will be given access permission and guidance to aid reproducibility and outcomes assessment. A Frequently Asked Questions about the data and access has been created at: https://ncats.nih.gov/n3c/about/program-faq 
The data model is OMOP 5.3.1, specifications are posted at: https://ncats.nih.gov/files/OMOP_CDM_COVID.pdf 

https://ncats.nih.gov/n3c

## Acknowledgements

The analyses described in this publication were conducted with data or tools accessed through the NCATS N3C Data Enclave covid.cd2h.org/enclave and supported by NCATS U24 TR002306. Halie M. Rando was supported by The Gordon and Betty Moore Foundation (GBMF 4552) and the National Human Genome Research Institute (R01 HG010067); Halie M. Rando supported by The Gordon and Betty Moore Foundation (GBMF 4552) and the National Human Genome Research Institute (R01 HG010067); Tellen D. Bennett supported by NIH UL1TR002535 03S2 and NIH UL1TR002535; James Brian Byrd supported by NIH grant K23HL128909 protected Dr. Byrd’s time to participate.; Christopher G. Chute supported by U24 TR002306; Rachel Deer supported by UTMB CTSA, 2P30AG024832-16 (PI: Volpi). This research was possible because of the patients whose information is included within the data from participating organizations (covid.cd2h.org/dtas) and scientists who have contributed to the on-going development of this community resource.

The project described was supported by the National Institute of General Medical Sciences, 5U54GM104942-04. The content is solely the responsibility of the authors and does not necessarily represent the official views of the NIH.

Data partners include but are not limited to the following: Carilion Clinic (UL1TR003015-02S2: Provision of Clinical Data to Support a Nationwide COVID-19 Cohort Collaborative); George Washington Children’s Research Institute (UL1TR001876: Clinical and Translational Science Institute at Children’s National); Duke University (UL1TR002553: Duke CTSA); Johns Hopkins University (UL1TR003098: Johns Hopkins Institute for Clinical and Translational Research); Mayo Clinic Rochester (UL1TR002377: Mayo Clinic Center for Clinical and Translational Science); Medical University of South Carolina (UL1TR001450: South Carolina Clinical & Translational Research Institute SCTR); Penn State Health Milton S. Hershey Medical Center (UL1TR002014: Penn State Clinical and Translational Science Institute); Rush University Medical Center (UL1TR002389: Institute for Translational Medicine); Stony Brook University; The Ohio State University (UL1TR002733: The OSU Center for Clinical and Translational Science: Advancing Today’s Discoveries to Improve Health); Tufts University Boston (UL1TR002544-03S4: Tufts Clinical and Translational Science Institute N3C Supplement); University of Massachusetts Medical School Worcester (UL1TR001453: University of Massachusetts Center for Clinical and Translational Science); University of Alabama at Birmingham (UL1TR003096: Center for Clinical and Translational Science); University of Arkansas for Medical Sciences (UL1TR003107: UAMS Translational Research Institute); The University of Chicago (UL1TR002389: ITM 2.0: Advancing Translational Science in Metropolitan Chicago); University of Colorado Denver (UL1TR002535-03S2: CCTSI Participation in the National COVID Cohort Collaborative N3C); University of Illinois at Chicago (UL1TR002003: Clinical and Translational Science Award); The University of Iowa (UL1TR002537: The University of Iowa Clinical and Translational Science Award); University of Kentucky (UL1TR001998-04S1: Kentucky Center for Clinical and Translational Science); University of Miami (UL1TR002736: Miami Clinical and Translational Science Institute); The University of Michigan at Ann Arbor (UL1TR002240: Michigan Institute for Clinical and Health Research); University of Minnesota (UL1TR002494: University of Minnesota Clinical and Translational Science Institute); University of Nebraska Lincoln (U54GM115458: University of Nebraska Center for Clinical & Translational Research); University of North Carolina at Chapel Hill (UL1TR002489: ICEES+ COVID-19 Open Infrastructure to Democratize and Accelerate Cross-Institutional Clinical Data Sharing and Research); University of Southern California (UL1TR001855: Southern California Clinical and Translational Institute); The University of Texas Medical Branch at Galveston (UL1TR001439: UTMB Clinical and Translational Science Award); The University of Utah (UL1TR002538-03S3: Infrastructure Support for Participation in the N3C Data Repository); University of Washington (UL1TR002319: Institute of Translational Health Sciences); University of Wisconsin-Madison (UL1TR002373: Institutional Clinical AND Translational Science Award); University of Virginia (UL1TR003015-02S2: Provision of Clinical Data to Support a Nationwide COVID-19 Cohort Collaborative); Virginia Commonwealth University (UL1TR002649-03S3: N3C & All of Us Research Program Collaborative Project); Wake Forest University Health Sciences (UL1TR001420: Wake Forest Clinical and Translational Science Award); Washington University in St. Louis (UL1TR002345: Washington University Institute of Clinical Translational Sciences); West Virginia University (U54GM104942: West Virginia Clinical and Translational Science Institute)

## Contributions

Contributions are organized according to contribution roles as follow:

- **data curation**: Halie M. Rando, Tiffany J. Callahan, Christopher G. Chute, Hannah Davis, Rachel Deer, Feifan Liu, Julie A. McMurry, Emily R. Pfaff, Rose Relevo, Peter N. Robinson, Melissa A. Haendel
- **data integration**: Tiffany J. Callahan, Christopher G. Chute, Feifan Liu, Emily R. Pfaff, Peter N. Robinson, Melissa A. Haendel
- **data quality assurance**: Christopher G. Chute, Emily R. Pfaff
- **data visualization**: Julie A. McMurry, Peter N. Robinson
- **manuscript review and editing**: Rachel Deer
- **clinical subject matter expertise**: Tellen D. Bennett, James Brian Byrd, Hannah Davis, Rachel Deer (patient perspective), Joel Gagnier, Farrukh M Koraishy, Joel H. Saltz
- **manuscript drafting**: Halie M. Rando, Tiffany J. Callahan, Christopher G. Chute, Rachel Deer, Farrukh M Koraishy, Feifan Liu, Julie A. McMurry, Emily R. Pfaff, Justin T. Reese, Rose Relevo, Peter N. Robinson, Joel H. Saltz, Anthony Solomonides, Melissa A. Haendel
- **project management**: Julie A. McMurry, Melissa A. Haendel
- **biological subject matter expertise**: Halie M. Rando, Justin T. Reese, Joel H. Saltz, Melissa A. Haendel
- **funding acquisition**: Christopher G. Chute
- **database / information systems admin**: Rose Relevo
- **clinical data model expertise**: Christopher G. Chute, Feifan Liu, Emily R. Pfaff, Peter N. Robinson
- **N3C Phenotype definition**: Carolyn Bramante, Christopher G. Chute, Farrukh M Koraishy, Emily R. Pfaff, Peter N. Robinson, Melissa A. Haendel
- **statistical analysis**: Justin T. Reese
- **governance**: Christopher G. Chute, Melissa A. Haendel
- **project evaluation**: Christopher G. Chute, Julie A. McMurry
- **critical revision of the manuscript for important intellectual content**: Halie M. Rando, Tellen D. Bennett, James Brian Byrd, Carolyn Bramante, Hannah Davis, Joel Gagnier, Farrukh M Koraishy, Feifan Liu, Julie A. McMurry, Richard A. Moffitt, Peter N. Robinson, Joel H. Saltz, Melissa A. Haendel

## Data Sharing

The N3C Data Enclave (covid.cd2h.org/enclave) houses fully reproducible, transparent, and broadly available limited and de-identified datasets (HIPAA definitions: https://www.hhs.gov/hipaa/for-professionals/privacy/specialtopics/de-identification/index.html).Data is accessible by investigators at institutions that have signed a Data Use Agreement with NIH who have taken human subjects and security training and attest to the N3C User Code of Conduct. Investigators wishing to access the limited dataset must also supply an institutional IRB protocol. All requests for data access are reviewed by the NIH Data Access Committee. A full description of the N3C Enclave governance has been published;^193^ information about how to apply for access is available on the NCATS website: ncats.nih.gov/n3c/about/applying-for-access. Reviewers and health authorities will be given access permission and guidance to aid reproducibility and outcomes assessment. A Frequently Asked Questions about the data and access has been created at: ncats.nih.gov/n3c/about/program-faq

The data model is OMOP 5.3.1, specifications are posted at: https://ncats.nih.gov/files/OMOP_CDM_COVID.pdf

## References

1 Puelles VG, Lütgehetmann M, Lindenmeyer MT, et al. Multiorgan and Renal Tropism of SARS-CoV-2. N Engl J Med 2020; 383: 590–2.

2 Gavriatopoulou M, Korompoki E, Fotiou D, et al. Organ-specific manifestations of COVID-19 infection. Clin Exp Med 2020; 20: 493–506.

3 Disser NP, De Micheli AJ, Schonk MM, et al. Musculoskeletal Consequences of COVID-19. J Bone Joint Surg Am 2020; 102: 1197–204.

4 Abdullahi A, Candan SA, Abba MA, et al. Neurological and Musculoskeletal Features of COVID-19: A Systematic Review and Meta-Analysis. Front Neurol 2020; 11: 687.

5 Ferrandi PJ, Alway SE, Mohamed JS. The interaction between SARS-CoV-2 and ACE2 may have consequences for skeletal muscle viral susceptibility and myopathies. J Appl Physiol 2020; 129: 864–7.

6 Harrison AG, Lin T, Wang P. Mechanisms of SARS-CoV-2 Transmission and Pathogenesis. Trends Immunol 2020; 41: 1100–15.

7 Cucinotta D, Vanelli M. WHO Declares COVID-19 a Pandemic. Acta Biomed 2020; 91: 157–60.

8 Gao Z, Xu Y, Sun C, et al. A systematic review of asymptomatic infections with COVID-19. J Microbiol Immunol Infect 2021; 54: 12–6.

9 Sykes DL, Holdsworth L, Jawad N, Gunasekera P, Morice AH, Crooks MG. Post-COVID-19 Symptom Burden: What is Long-COVID and How Should We Manage It? Lung 2021; published online Feb 11. DOI:10.1007/s00408-021-00423-z.

10 Gavinio R. Rev Med Suisse 2021; 17: 322.

11 Zhang J, Xu J, Zhou S, et al. The characteristics of 527 discharged COVID-19 patients undergoing long-term follow-up in China. Int J Infect Dis 2021; 104: 685–92.

12 Kedor C, Freitag H, Meyer-Arndt L, et al. Chronic COVID-19 Syndrome and Chronic Fatigue Syndrome (ME/CFS) following the first pandemic wave in Germany – a first analysis of a prospective observational study. bioRxiv. 2021; published online Feb 8. DOI:10.1101/2021.02.06.21249256.

13 Davis HE, Assaf GS, McCorkell L, et al. Characterizing Long COVID in an international cohort: 7 months of symptoms and their impact. bioRxiv. 2020; published online Dec 26. DOI:10.1101/2020.12.24.20248802.

14 Huang C, Huang L, Wang Y, et al. 6-month consequences of COVID-19 in patients discharged from hospital: a cohort study. Lancet 2021; 397: 220–32.

15 Kirby T. COVID-19 survivor experiencing long-term symptoms. Lancet Respir Med 2021; published online Feb 12. DOI:10.1016/S2213-2600(21)00092-8.

16 Edwards E, Dunn L, Gosk S, et al. Inside ‘post-Covid’ clinics: How specialized centers are trying to treat long-haulers. NBC News. 2021; published online March 1. https://www.nbcnews.com/health/health-news/inside-post-covid-clinics-how-specialized-centers-are-trying-treat-n1258879 (accessed March 4, 2021).

17 White PD, Thomas JM, Kangro HO, et al. Predictions and associations of fatigue syndromes and mood disorders that occur after infectious mononucleosis. Lancet 2001; 358: 1946–54.

18 Hickie I, Davenport T, Wakefield D, et al. Post-infective and chronic fatigue syndromes precipitated by viral and non-viral pathogens: prospective cohort study. BMJ 2006; 333: 575.

19 Wensaas K-A, Langeland N, Hanevik K, Mørch K, Eide GE, Rortveit G. Irritable bowel syndrome and chronic fatigue 3 years after acute giardiasis: historic cohort study. Gut 2012; 61: 214–9.

20 Bansal AS, Bradley AS, Bishop KN, Kiani-Alikhan S, Ford B. Chronic fatigue syndrome, the immune system and viral infection. Brain Behav Immun 2012; 26: 24–31.

21 Moldofsky H, Patcai J. Chronic widespread musculoskeletal pain, fatigue, depression and disordered sleep in chronic post-SARS syndrome; a case-controlled study. BMC Neurol 2011; 11: 37.

22 Lam MH-B, Wing Y-K, Yu MW-M, et al. Mental morbidities and chronic fatigue in severe acute respiratory syndrome survivors: long-term follow-up. Arch Intern Med 2009; 169: 2142–7.

23 Lee SH, Shin H-S, Park HY, et al. Depression as a Mediator of Chronic Fatigue and Post-Traumatic Stress Symptoms in Middle East Respiratory Syndrome Survivors. Psychiatry Investig 2019; 16: 59–64.

24 Ong K-C, Ng AW-K, Lee LS-U, et al. 1-year pulmonary function and health status in survivors of severe acute respiratory syndrome. Chest 2005; 128: 1393–400.

25 Ngai JC, Ko FW, Ng SS, To K-W, Tong M, Hui DS. The long-term impact of severe acute respiratory syndrome on pulmonary function, exercise capacity and health status. Respirology 2010; 15: 543–50.

26 Hui DS, Ko FW, Chan DP, et al. The long-term impact of severe acute respiratory syndrome (SARS) on pulmonary function, exercise capacity, and quality of life in a cohort of survivors. Chest 2005; 128: 148S.

27 Zhang P, Li J, Liu H, et al. Long-term bone and lung consequences associated with hospital-acquired severe acute respiratory syndrome: a 15-year follow-up from a prospective cohort study. Bone Res 2020; 8: 8.

28 Wu Q, Zhou L, Sun X, et al. Altered Lipid Metabolism in Recovered SARS Patients Twelve Years after Infection. Sci Rep 2017; 7: 9110.

29 Yin C-H, Wang C, Wen Y, et al. [Prospective 2-year clinical study of patients with positive IgG-antibodies after recovering from severe acute respiratory syndrome]. Zhongguo Wei Zhong Bing Ji Jiu Yi Xue 2005; 17: 740–2.

30 Liu Y-X, Ye Y-P, Zhang P, et al. [Changes in pulmonary function in SARS patients during the three-year convalescent period]. Zhongguo Wei Zhong Bing Ji Jiu Yi Xue 2007; 19: 536–8.

31 Il Jun K, Park WB, Kim G, et al. Long-term Respiratory Complication in Patients with Middle East Respiratory Syndrome: 1-year Follow-up After the 2015 Outbreak in South Korea. Open Forum Infect Dis 2017; 4: S577–S577.

32 Cirulli ET, Schiabor Barrett KM, Riffle S, et al. Long-term COVID-19 symptoms in a large unselected population. bioRxiv. 2020; published online Oct 11. DOI:10.1101/2020.10.07.20208702.

33 Lopez-Leon S, Wegman-Ostrosky T, Perelman C, et al. More than 50 Long-term effects of COVID-19: a systematic review and meta-analysis. medRxiv 2021; published online Jan 30. DOI:10.1101/2021.01.27.21250617.

34 Callard F, Perego E. How and why patients made Long Covid. Soc Sci Med 2021; 268: 113426.

35 Perego E, Callard F, Stras L, Melville-Jóhannesson B, Pope R, Alwan NA. Why the Patient-Made Term ‘Long Covid’ is needed. Wellcome Open Res 2020; 5: 224.

36 Fiona Lowenstein And. Opinion. The New York Times. 2021; published online March 17. https://www.nytimes.com/2021/03/17/opinion/long-covid.html (accessed March 19, 2021).

37 Greenhalgh T, Knight M, A’Court C, Buxton M, Husain L. Management of post-acute covid-19 in primary care. BMJ 2020; 370: m3026.

38 Gomersall CD, Chan DP, Leung P, Joynt GM, Hui DS. Long term outcome of acute respiratory distress syndrome caused by severe acute respiratory syndrome (SARS): an observational study. Crit Care Resusc 2006; published online Dec. https://search.informit.com.au/documentSummary;dn=516810758621590;res=IELHEA (accessed March 2, 2021).

39 Rawal G, Yadav S, Kumar R. Post-intensive Care Syndrome: an Overview. J Transl Int Med 2017; 5: 90–2.

40 Lever J, Altman RB. Analyzing the vast coronavirus literature with CoronaCentral. bioRxiv 2020; published online Dec 22. DOI:10.1101/2020.12.21.423860.

41 Mahase E. Covid-19: What do we know about ‘long covid’? BMJ 2020; 370: m2815.

42 Datta SD, Talwar A, Lee JT. A Proposed Framework and Timeline of the Spectrum of Disease Due to SARS-CoV-2 Infection: Illness Beyond Acute Infection and Public Health Implications. JAMA 2020; 324: 2251–2.

43 Vogel TP, Top KA, Karatzios C, et al. Multisystem inflammatory syndrome in children and adults (MIS-C/A): Case definition & guidelines for data collection, analysis, and presentation of immunization safety data. Vaccine 2021; published online Feb 24. DOI:10.1016/j.vaccine.2021.01.054.

44 Sivan M, Taylor S. NICE guideline on long covid. BMJ 2020; 371: m4938.

45 Amenta EM, Spallone A, Rodriguez-Barradas MC, El Sahly HM, Atmar RL, Kulkarni PA. Postacute COVID-19: An Overview and Approach to Classification. Open Forum Infect Dis 2020; 7: ofaa509.

46 Ladds E, Rushforth A, Wieringa S, et al. Persistent symptoms after Covid-19: qualitative study of 114 ‘long Covid’ patients and draft quality principles for services. BMC Health Serv Res 2020; 20: 1144.

47 Ludvigsson JF. Case report and systematic review suggest that children may experience similar long-term effects to adults after clinical COVID-19. Acta Paediatr 2021; 110: 914–21.

48 Flu-pro©. 2020; published online April 13. https://www.evidera.com/flu-pro/ (accessed March 18, 2021).

49 EQ-5D-5L – EQ-5D. https://euroqol.org/eq-5d-instruments/eq-5d-5l-about/ (accessed March 18, 2021).

50 Williams N. The MRC breathlessness scale. Occup Med 2017; 67: 496–7.

51 Köhler S, Gargano M, Matentzoglu N, et al. The Human Phenotype Ontology in 2021. Nucleic Acids Res 2021; 49: D1207–17.

52 Goërtz YMJ, Van Herck M, Delbressine JM, et al. Persistent symptoms 3 months after a SARS-CoV-2 infection: the post-COVID-19 syndrome? ERJ Open Res 2020; 6. DOI:10.1183/23120541.00542-2020.

53 Ho DE, Imai K, King G, Stuart EA. MatchIt: Nonparametric Preprocessing for Parametric Causal Inference. J Stat Softw 2011; 42. DOI:10.18637/jss.v042.i08.

54 callahantiff. callahantiff/OMOP2OBO. https://github.com/callahantiff/OMOP2OBO (accessed March 10, 2021).

55 Athena. https://athena.ohdsi.org/search-terms/start (accessed March 10, 2021).

56 Zhao Y-M, Shang Y-M, Song W-B, et al. Follow-up study of the pulmonary function and related physiological characteristics of COVID-19 survivors three months after recovery. EClinicalMedicine 2020; 25: 100463.

57 Liu D, Zhang W, Pan F, et al. The pulmonary sequalae in discharged patients with COVID-19: a short-term observational study. Respir Res 2020; 21: 125.

58 Townsend L, Dyer AH, Jones K, et al. Persistent fatigue following SARS-CoV-2 infection is common and independent of severity of initial infection. PLoS One 2020; 15: e0240784.

59 Tenforde MW, Kim SS, Lindsell CJ, et al. Symptom Duration and Risk Factors for Delayed Return to Usual Health Among Outpatients with COVID-19 in a Multistate Health Care Systems Network — United States, March–June 2020. MMWR. Morbidity and Mortality Weekly Report. 2020; 69: 993–8.

60 Dani M, Dirksen A, Taraborrelli P, et al. Autonomic dysfunction in ‘long COVID’: rationale, physiology and management strategies. Clin Med 2021; 21: e63–7.

61 Dennis A, Wamil M, Kapur S, et al. Multi-organ impairment in low-risk individuals with long COVID. bioRxiv. 2020; published online Oct 16. DOI:10.1101/2020.10.14.20212555.

62 Doykov I, Hällqvist J, Gilmour KC, Grandjean L, Mills K, Heywood WE. ‘The long tail of Covid-19’ -The detection of a prolonged inflammatory response after a SARS-CoV-2 infection in asymptomatic and mildly affected patients. F1000Res 2020; 9: 1349.

63 Mandal S, Barnett J, Brill SE, et al. ‘Long-COVID’: a cross-sectional study of persisting symptoms, biomarker and imaging abnormalities following hospitalisation for COVID-19. Thorax 2020; published online Nov 10. DOI:10.1136/thoraxjnl-2020-215818.

64 McMahon DE, Gallman AE, Hruza GJ, et al. Long COVID in the skin: a registry analysis of COVID-19 dermatological duration. Lancet Infect Dis 2021; 21: 313–4.

65 Nehme M, Braillard O, Alcoba G, et al. COVID-19 Symptoms: Longitudinal Evolution and Persistence in Outpatient Settings. Ann Intern Med 2020; published online Dec 8. DOI:10.7326/M20-5926.

66 Petersen MS, Kristiansen MF, Hanusson KD, et al. Long COVID in the Faroe Islands - a longitudinal study among non-hospitalized patients. Clin Infect Dis 2020; published online Nov 30. DOI:10.1093/cid/ciaa1792.

67 Raahimi MM, Kane A, Moore CE, Alareed AW. Late onset of Guillain-Barré syndrome following SARS-CoV-2 infection: part of ‘long COVID-19 syndrome’? BMJ Case Rep 2021; 14. DOI:10.1136/bcr-2020-240178.

68 Sudre CH, Murray B, Varsavsky T, et al. Attributes and predictors of Long-COVID: analysis of COVID cases and their symptoms collected by the Covid Symptoms Study App. bioRxiv. 2020; published online Oct 21. DOI:10.1101/2020.10.19.20214494.

69 Zampogna E, Migliori GB, Centis R, et al. Functional impairment during post-acute COVID-19 phase: Preliminary finding in 56 patients. Pulmonology 2021; published online Jan 6. DOI:10.1016/j.pulmoe.2020.12.008.

70 Gregorova M, Morse D, Brignoli T, et al. Post-acute COVID-19 associated with evidence of bystander T-cell activation and a recurring antibiotic-resistant bacterial pneumonia. Elife 2020; 9. DOI:10.7554/eLife.63430.

71 Walsh-Messinger J, Manis H, Vrabec A, et al. The Kids Are Not Alright: A Preliminary Report of Post-COVID Syndrome in University Students. medRxiv 2020; published online Nov 29. DOI:10.1101/2020.11.24.20238261.

72 Moreno-Pérez O, Merino E, Leon-Ramirez J-M, et al. Post-acute COVID-19 syndrome. Incidence and risk factors: A Mediterranean cohort study. J Infect 2021; published online Jan 12. DOI:10.1016/j.jinf.2021.01.004.

73 Arnold DT, Hamilton FW, Milne A, et al. Patient outcomes after hospitalisation with COVID-19 and implications for follow-up: results from a prospective UK cohort. Thorax 2020; published online Dec 3. DOI:10.1136/thoraxjnl-2020-216086.

74 Blair PW, Brown DM, Jang M, et al. The Clinical Course of COVID-19 in the Outpatient Setting: A Prospective Cohort Study. Open Forum Infect Dis 2021; 8: ofab007.

75 Carfì A, Bernabei R, Landi F, Gemelli Against COVID-19 Post-Acute Care Study Group. Persistent Symptoms in Patients After Acute COVID-19. JAMA 2020; 324: 603–5.

76 Carvalho-Schneider C, Laurent E, Lemaignen A, et al. Follow-up of adults with noncritical COVID-19 two months after symptom onset. Clin Microbiol Infect 2021; 27: 258–63.

77 Chopra V, Flanders SA, O’Malley M, Malani AN, Prescott HC. Sixty-Day Outcomes Among Patients Hospitalized With COVID-19. Ann Intern Med 2020; published online Nov 11. DOI:10.7326/M20-5661.

78 Ding H, Yin S, Cheng Y, Cai Y, Huang W, Deng W. Neurologic manifestations of nonhospitalized patients with COVID-19 in Wuhan, China. MedComm 2020; 1: 253–6.

79 Garrigues E, Janvier P, Kherabi Y, et al. Post-discharge persistent symptoms and health-related quality of life after hospitalization for COVID-19. J Infect 2020; 81: e4–6.

80 Manckoundia P, Franon E. Is Persistent Thick Copious Mucus a Long-Term Symptom of COVID-19? Eur J Case Rep Intern Med 2020; 7: 002145.

81 Poyraz BÇ, Poyraz CA, Olgun Y, et al. Psychiatric morbidity and protracted symptoms after COVID-19. Psychiatry Res 2021; 295: 113604.

82 Varghese J, Sandmann S, Vollenberg R, et al. Follow up of COVID-19 features in recovered adults without comorbidities– persistent symptoms and lab-abnormalities. 2020; published online Nov 30. DOI:10.21203/rs.3.rs-116030/v1.

83 Bakhoum MF, Ritter M, Garg AK, Chan AX, Bakhoum CY, Smith DM. Subclinical ocular inflammation in persons recovered from ambulatory COVID-19. bioRxiv. 2020; published online Sept 23. DOI:10.1101/2020.09.22.20128140.

84 Brito D, Meester S, Yanamala N, et al. High Prevalence of Pericardial Involvement in College Student Athletes Recovering From COVID-19. JACC Cardiovasc Imaging 2021; 14: 541–55.

85 Galal I, Hussein AARM, Amin MT, et al. Determinants of Persistent Post COVID-19 symptoms: Value of a Novel COVID-19 symptoms score. bioRxiv. 2020; published online Nov 12. DOI:10.1101/2020.11.11.20230052.

86 Lu Y, Li X, Geng D, et al. Cerebral Micro-Structural Changes in COVID-19 Patients - An MRI-based 3-month Follow-up Study. EClinicalMedicine 2020; 25: 100484.

87 Puntmann VO, Carerj ML, Wieters I, et al. Outcomes of Cardiovascular Magnetic Resonance Imaging in Patients Recently Recovered From Coronavirus Disease 2019 (COVID-19). JAMA Cardiol 2020; 5: 1265–73.

88 Wong AW, Shah AS, Johnston JC, Carlsten C, Ryerson CJ. Patient-reported outcome measures after COVID-19: a prospective cohort study. Eur Respir J 2020; 56. DOI:10.1183/13993003.03276-2020.

89 Al-Jahdhami I, Al-Naamani K, Al-Mawali A. The Post-acute COVID-19 Syndrome (Long COVID). Oman Med J 2021; 36: e220.

90 Corsini Campioli C, Cano Cevallos E, Assi M, Patel R, Binnicker MJ, O’Horo JC. Clinical predictors and timing of cessation of viral RNA shedding in patients with COVID-19. J Clin Virol 2020; 130: 104577.

91 Zhou F, Yu T, Du R, et al. Clinical course and risk factors for mortality of adult inpatients with COVID-19 in Wuhan, China: a retrospective cohort study. Lancet 2020; 395: 1054–62.

92 Halpin SJ, McIvor C, Whyatt G, et al. Postdischarge symptoms and rehabilitation needs in survivors of COVID-19 infection: A cross-sectional evaluation. J Med Virol 2021; 93: 1013–22.

93 Xiong Q, Xu M, Li J, et al. Clinical sequelae of COVID-19 survivors in Wuhan, China: a single-centre longitudinal study. Clin Microbiol Infect 2021; 27: 89–95.

94 Liu C, Ye L, Xia R, et al. Chest Computed Tomography and Clinical Follow-Up of Discharged Patients with COVID-19 in Wenzhou City, Zhejiang, China. Ann Am Thorac Soc 2020; 17: 1231–7.

95 Liang L, Yang B, Jiang N, et al. Three-month Follow-up Study of Survivors of Coronavirus Disease 2019 after Discharge. J Korean Med Sci 2020; 35: e418.

96 Bellan M, Soddu D, Balbo PE, et al. Respiratory and Psychophysical Sequelae Among Patients With COVID-19 Four Months After Hospital Discharge. JAMA Netw Open 2021; 4: e2036142.

97 Logue JK, Franko NM, McCulloch DJ, et al. Sequelae in Adults at 6 Months After COVID-19 Infection. JAMA Netw Open 2021; 4: e210830.

98 Rando HM, MacLean AL, Lee AJ, et al. Pathogenesis, Symptomatology, and Transmission of SARS-CoV-2 through analysis of Viral Genomics and Structure. ArXiv 2021; published online Feb 1. https://www.ncbi.nlm.nih.gov/pubmed/33594340.

99 Wölfel R, Corman VM, Guggemos W, et al. Virological assessment of hospitalized patients with COVID-2019. Nature 2020; 581: 465–9.

100 Arons MM, Hatfield KM, Reddy SC, et al. Presymptomatic SARS-CoV-2 Infections and Transmission in a Skilled Nursing Facility. N Engl J Med 2020; 382: 2081–90.

101 Bullard J, Dust K, Funk D, et al. Predicting infectious SARS-CoV-2 from diagnostic samples. Clin Infect Dis 2020; published online May 22. DOI:10.1093/cid/ciaa638.

102 CDC. Interim guidance on duration of isolation and precautions for adults with COVID-19. 2021; published online Feb 14. https://www.cdc.gov/coronavirus/2019-ncov/hcp/duration-isolation.html (accessed Feb 25, 2021).

103 Mei Q, Li J, Du R, Yuan X, Li M, Li J. Assessment of patients who tested positive for COVID-19 after recovery. Lancet Infect Dis 2020; 20: 1004–5.

104 Lu J, Peng J, Xiong Q, et al. Clinical, immunological and virological characterization of COVID-19 patients that test re-positive for SARS-CoV-2 by RT-PCR. EBioMedicine 2020; 59: 102960.

105 KDCA. KDCA. https://www.cdc.go.kr/board/board.es?mid=a30402000000&bid=0030&act=view&list_no=367267&nPage=1external%20icon (accessed Feb 25, 2021).

106 Gaebler C, Wang Z, Lorenzi JCC, et al. Evolution of antibody immunity to SARS-CoV-2. Nature 2021; published online Jan 18. DOI:10.1038/s41586-021-03207-w.

107 Confronting the pathophysiology of long covid -The BMJ. 2020; published online Dec 9. https://blogs.bmj.com/bmj/2020/12/09/confronting-the-pathophysiology-of-long-covid/ (accessed March 8, 2021).

108 Kanji JN, Zelyas N, MacDonald C, et al. False negative rate of COVID-19 PCR testing: a discordant testing analysis. Virol J 2021; 18: 13.

109 Woloshin S, Patel N, Kesselheim AS. False Negative Tests for SARS-CoV-2 Infection -Challenges and Implications. N Engl J Med 2020; 383: e38.

110 Jia X, Xiao L, Liu Y. False negative RT-PCR and false positive antibody tests-Concern and solutions in the diagnosis of COVID-19. J Infect 2020; published online Oct 8. DOI:10.1016/j.jinf.2020.10.007.

111 Watson J, Richter A, Deeks J. Testing for SARS-CoV-2 antibodies. BMJ 2020; 370: m3325.

112 Allen WE, Altae-Tran H, Briggs J, et al. Population-scale Longitudinal Mapping of COVID-19 Symptoms, Behavior, and Testing Identifies Contributors to Continued Disease Spread in the United States. medRxiv 2020; published online June 11. DOI:10.1101/2020.06.09.20126813.

113 Needham DM, Davidson J, Cohen H, et al. Improving long-term outcomes after discharge from intensive care unit: report from a stakeholders’ conference. Crit Care Med 2012; 40: 502–9.

114 Dufort EM, Koumans EH, Chow EJ, et al. Multisystem Inflammatory Syndrome in Children in New York State. N Engl J Med 2020; 383: 347–58.

115 Weatherhead JE, Clark E, Vogel TP, Atmar RL, Kulkarni PA. Inflammatory syndromes associated with SARS-CoV-2 infection: dysregulation of the immune response across the age spectrum. J Clin Invest 2020; 130: 6194–7.

116 Wostyn P. COVID-19 and chronic fatigue syndrome: Is the worst yet to come? Med Hypotheses 2021; 146: 110469.

117 Fraser DD, Patterson EK, Slessarev M, et al. Endothelial Injury and Glycocalyx Degradation in Critically Ill Coronavirus Disease 2019 Patients: Implications for Microvascular Platelet Aggregation. Crit Care Explor 2020; 2: e0194.

118 Rovas A, Osiaevi I, Buscher K, et al. Microvascular dysfunction in COVID-19: the MYSTIC study. Angiogenesis 2021; 24: 145–57.

119 Yamaoka-Tojo M. Endothelial glycocalyx damage as a systemic inflammatory microvascular endotheliopathy in COVID-19. Biomed J 2020; 43: 399–413.

120 Hsu RK, Hsu C-Y. The Role of Acute Kidney Injury in Chronic Kidney Disease. Semin Nephrol 2016; 36: 283–92.

121 Chawla LS, Eggers PW, Star RA, Kimmel PL. Acute kidney injury and chronic kidney disease as interconnected syndromes. N Engl J Med 2014; 371: 58–66.

122 Wang S, Zhou X, Zhang T, Wang Z. The need for urogenital tract monitoring in COVID-19. Nat Rev Urol 2020; 17: 314–5.

123 Fanelli V, Fiorentino M, Cantaluppi V, et al. Acute kidney injury in SARS-CoV-2 infected patients. Crit. Care. 2020; 24: 155.

124 Hayden MR, Yang Y, Habibi J, Bagree SV, Sowers JR. Pericytopathy: oxidative stress and impaired cellular longevity in the pancreas and skeletal muscle in metabolic syndrome and type 2 diabetes. Oxid Med Cell Longev 2010; 3: 290–303.

125 Gansevoort RT, Correa-Rotter R, Hemmelgarn BR, et al. Chronic kidney disease and cardiovascular risk: epidemiology, mechanisms, and prevention. Lancet 2013; 382: 339–52.

126 Tonelli M, Wiebe N, Culleton B, et al. Chronic kidney disease and mortality risk: a systematic review. J Am Soc Nephrol 2006; 17: 2034–47.

127 Hultström M, Lipcsey M, Wallin E, Larsson I-M, Larsson A, Frithiof R. Severe acute kidney injury associated with progression of chronic kidney disease after critical COVID-19. Crit Care 2021; 25: 37.

128 Coca SG, Singanamala S, Parikh CR. Chronic kidney disease after acute kidney injury: a systematic review and meta-analysis. Kidney Int 2012; 81: 442–8.

129 Ng JH, Hirsch JS, Hazzan A, et al. Outcomes Among Patients Hospitalized With COVID-19 and Acute Kidney Injury. Am J Kidney Dis 2021; 77: 204–15.e1.

130 Chan L, Chaudhary K, Saha A, et al. Acute Kidney Injury in Hospitalized Patients with COVID-19. medRxiv 2020; published online May 8. DOI:10.1101/2020.05.04.20090944.

131 Oronsky B, Larson C, Hammond TC, et al. A Review of Persistent Post-COVID Syndrome (PPCS). Clin Rev Allergy Immunol 2021; published online Feb 20. DOI:10.1007/s12016-021-08848-3.

132 Mahammedi A, Saba L, Vagal A, et al. Imaging of Neurologic Disease in Hospitalized Patients with COVID-19: An Italian Multicenter Retrospective Observational Study. Radiology 2020; 297: E270–3.

133 Klironomos S, Tzortzakakis A, Kits A, et al. Nervous System Involvement in Coronavirus Disease 2019: Results from a Retrospective Consecutive Neuroimaging Cohort. Radiology 2020; 297: E324–34.

134 Kremer S, Lersy F, Anheim M, et al. Neurologic and neuroimaging findings in patients with COVID-19: A retrospective multicenter study. Neurology 2020; 95: e1868–82.

135 Kremer S, Lersy F, de Sèze J, et al. Brain MRI Findings in Severe COVID-19: A Retrospective Observational Study. Radiology 2020; 297: E242–51.

136 Chetrit A, Lechien JR, Ammar A, et al. Magnetic resonance imaging of COVID-19 anosmic patients reveals abnormalities of the olfactory bulb: Preliminary prospective study. J. Infect. 2020; 81: 816–46.

137 Guedj E, Campion JY, Dudouet P, et al. 18F-FDG brain PET hypometabolism in patients with long COVID. Eur J Nucl Med Mol Imaging 2021; published online Jan 26. DOI:10.1007/s00259-021-05215-4.

138 Yong SJ. Persistent Brainstem Dysfunction in Long-COVID: A Hypothesis. ACS Chem Neurosci 2021; 12: 573–80.

139 Fidahic M, Nujic D, Runjic R, et al. Research methodology and characteristics of journal articles with original data, preprint articles and registered clinical trial protocols about COVID-19. BMC Med Res Methodol 2020; 20: 161.

140 Serrano GE, Walker JE, Arce R, et al. Mapping of SARS-CoV-2 Brain Invasion and Histopathology in COVID-19 Disease. medRxiv 2021; published online Feb 18. DOI:10.1101/2021.02.15.21251511.

141 Chaudhri I, Moffitt R, Taub E, et al. Association of Proteinuria and Hematuria with Acute Kidney Injury and Mortality in Hospitalized Patients with COVID-19. Kidney Blood Press Res 2020; 45: 1018–32.

142 Stony Brook COVID-19 Research Consortium. Geospatial Distribution and Predictors of Mortality in Hospitalized Patients With COVID-19: A Cohort Study. Open Forum Infect Dis 2020; 7: ofaa436.

143 Chu KH, Tsang WK, Tang CS, et al. Acute renal impairment in coronavirus-associated severe acute respiratory syndrome. Kidney Int 2005; 67: 698–705.

144 Bradley BT, Maioli H, Johnston R, et al. Histopathology and ultrastructural findings of fatal COVID-19 infections in Washington State: a case series. Lancet 2020; 396: 320–32.

145 Sharma P, Ng JH, Bijol V, Jhaveri KD, Wanchoo R. Pathology of COVID 19 associated acute kidney injury. Clin Kidney J 2021; published online Jan 24. DOI:10.1093/ckj/sfab003.

146 Su H, Yang M, Wan C, et al. Renal histopathological analysis of 26 postmortem findings of patients with COVID-19 in China. Kidney Int 2020; 98: 219–27.

147 Müller JA, Groß R, Conzelmann C, et al. SARS-CoV-2 infects and replicates in cells of the human endocrine and exocrine pancreas. Nat Metab 2021; 3: 149–65.

148 Mukerji SS, Solomon IH. What can we learn from brain autopsies in COVID-19? Neurosci Lett 2021; 742: 135528.

149 Nauen DW, Hooper JE, Stewart CM, Solomon IH. Assessing Brain Capillaries in Coronavirus Disease 2019. JAMA Neurol 2021; published online Feb 12. DOI:10.1001/jamaneurol.2021.0225.

150 Rapkiewicz AV, Mai X, Carsons SE, et al. Megakaryocytes and platelet-fibrin thrombi characterize multi-organ thrombosis at autopsy in COVID-19: A case series. EClinicalMedicine. 2020; 24: 100434.

151 Duarte-Neto AN, Monteiro RAA, da Silva LFF, et al. Pulmonary and systemic involvement in COVID-19 patients assessed with ultrasound-guided minimally invasive autopsy. Histopathology 2020; 77: 186–97.

152 Santoriello D, Khairallah P, Bomback AS, et al. Postmortem Kidney Pathology Findings in Patients with COVID-19. J Am Soc Nephrol 2020; 31: 2158–67.

153 Zijlstra JG, van Meurs M, Moser J. Post-Mortem Diagnostics in COVID-19 AKI, More Often but Timely. J. Am. Soc. Nephrol. 2021; 32: 255.

154 Bouquegneau A, Erpicum P, Grosch S, Habran L. COVID-19-associated nephropathy includes tubular necrosis and capillary congestion, with evidence of SARS-CoV-2 in the nephron. Kidney360 2021. https://kidney360.asnjournals.org/content/early/2021/02/12/KID.0006992020.abstract.

155 Sun S, Lin D, Operario D. Need for a population health approach to understand and address psychosocial consequences of COVID-19. Psychol Trauma 2020; 12: S25–7.

156 Dubey S, Biswas P, Ghosh R, et al. Psychosocial impact of COVID-19. Diabetes Metab Syndr 2020; 14: 779–88.

157 Pfefferbaum B, North CS. Mental Health and the Covid-19 Pandemic. N Engl J Med 2020; 383: 510–2.

158 Otu A, Charles CH, Yaya S. Mental health and psychosocial well-being during the COVID-19 pandemic: the invisible elephant in the room. Int J Ment Health Syst 2020; 14: 38.

159 Clauw DJ, Häuser W, Cohen SP, Fitzcharles M-A. Considering the potential for an increase in chronic pain after the COVID-19 pandemic. Pain 2020; 161: 1694–7.

160 Ashby NJS. Impact of the COVID-19 Pandemic on Unhealthy Eating in Populations with Obesity. Obesity 2020; 28: 1802–5.

161 Myers CA, Broyles ST. Fast Food Patronage and Obesity Prevalence During the COVID-19 Pandemic: An Alternative Explanation. Obesity. 2020; 28: 1796–7.

162 Mutz M, Gerke M. Sport and exercise in times of self-quarantine: How Germans changed their behaviour at the beginning of the Covid-19 pandemic. Int Rev Sociol Sport 2020; : 1012690220934335.

163 Volkow ND. Collision of the COVID-19 and Addiction Epidemics. Ann Intern Med 2020; 173: 61– 2.

164 Czeisler MÉ, Lane RI, Petrosky E, et al. Mental Health, Substance Use, and Suicidal Ideation During the COVID-19 Pandemic - United States, June 24-30, 2020. MMWR Morb Mortal Wkly Rep 2020; 69: 1049–57.

165 Zaami S, Marinelli E, Varì MR. New Trends of Substance Abuse During COVID-19 Pandemic: An International Perspective. Front Psychiatry 2020; 11: 700.

166 Lipsitch M, Swerdlow DL, Finelli L. Defining the Epidemiology of Covid-19 - Studies Needed. N Engl J Med 2020; 382: 1194–6.

167 Lythgoe MP, Middleton P. Ongoing Clinical Trials for the Management of the COVID-19 Pandemic. Trends Pharmacol Sci 2020; 41: 363–82.

168 COVID-19 Views - ClinicalTrials.gov. Views of COVID-19 Studies Listed on ClinicalTrials.gov (Beta). https://clinicaltrials.gov/ct2/covid_view (accessed March 9, 2021).

169 Kim AHJ, Sparks JA, Liew JW, et al. A Rush to Judgment? Rapid Reporting and Dissemination of Results and Its Consequences Regarding the Use of Hydroxychloroquine for COVID-19. Ann Intern Med 2020; 172: 819–21.

170 Jin X, Pang B, Zhang J, et al. Core Outcome Set for Clinical Trials on Coronavirus Disease 2019 (COS-COVID). Engineering 2020; 6: 1147–52.

171 Qiu R, Wei X, Zhao M, et al. Outcome reporting from protocols of clinical trials of Coronavirus Disease 2019 (COVID-19): a review. medRxiv 2020. https://www.medrxiv.org/content/10.1101/2020.03.04.20031401v1.abstract.

172 Wang H, Jin X, Pang B, Others. Analysis of clinical research scheme of traditional Chinese medicine intervention on COVID-19. Chin J Trad Chin Med 2020; : 1232–41.

173 Alexander PE, Debono VB, Mammen MJ, et al. COVID-19 coronavirus research has overall low methodological quality thus far: case in point for chloroquine/hydroxychloroquine. J Clin Epidemiol 2020; 123: 120–6.

174 Kirkham JJ, Gargon E, Clarke M, Williamson PR. Can a core outcome set improve the quality of systematic reviews?--a survey of the Co-ordinating Editors of Cochrane Review Groups. Trials 2013; 14: 21.

175 Williamson PR, Altman DG, Blazeby JM, et al. Developing core outcome sets for clinical trials: issues to consider. Trials 2012; 13: 132.

176 Clarke M. Standardising outcomes for clinical trials and systematic reviews. Trials 2007; 8: 39.

177 Boers M, Kirwan JR, Wells G, et al. Developing core outcome measurement sets for clinical trials: OMERACT filter 2.0. J Clin Epidemiol 2014; 67: 745–53.

178 Standards and Indicators | NICE. Quality standards set out the priority areas for quality improvement in health and social care. https://www.nice.org.uk/standards-and-indicators (accessed March 9, 2021).

179 Petzold T, Deckert S, Williamson PR, Schmitt J. Quality Measurement Recommendations Relevant to Clinical Guidelines in Germany and the United Kingdom: (What) Can We Learn From Each Other? Inquiry 2018; 55: 46958018761495.

180 Donabedian A. The quality of care: how can it be assessed? JAMA 1988; 260: 1743–8.

181 Boulkedid R, Abdoul H, Loustau M, Sibony O, Alberti C. Using and reporting the Delphi method for selecting healthcare quality indicators: a systematic review. PLoS One 2011; 6: e20476.

182 Porter ME, Others. A strategy for health care reform—toward a value-based system. N Engl J Med 2009; 361: 109–12.

183 Hurst L, Mahtani K, Pluddemann A, et al. Defining Value-based Healthcare in the NHS. 2019. http://www.cebm.net/wp-content/uploads/2019/04/Defining-Value-based-healthcare-in-the-NHS-Final4-1.pdf.

184 National Institute for Health and Care Excellence. Developing NICE Guidelines: The Manual. London: National Institute for Health and Care Excellence (NICE), 2015 https://www.ncbi.nlm.nih.gov/pubmed/26677490.

185 Austin JM, Kachalia A. The State of Health Care Quality Measurement in the Era of COVID-19: The Importance of Doing Better. JAMA 2020; 324: 333–4.

186 Qiu R, Zhao C, Liang T, et al. Core Outcome Set for Clinical Trials of COVID-19 Based on Traditional Chinese and Western Medicine. Front Pharmacol 2020; 11: 781.

187 Marshall JC, Murthy S, Diaz J, et al. A minimal common outcome measure set for COVID-19 clinical research. Lancet Infect Dis 2020; 20: e192–7.

188 Evangelidis N, Tong A, Howell M, et al. International survey to establish prioritized outcomes for trials in people with COVID-19. 2020. https://www.repository.cam.ac.uk/handle/1810/308249.

189 Arab-Zozani M, Hashemi F, Safari H, Yousefi M, Ameri H. Health-Related Quality of Life and its Associated Factors in COVID-19 Patients. Osong Public Health Res Perspect 2020; 11: 296– 302.

190 Tabacof L, Tosto-Mancuso J, Wood J, et al. Post-acute COVID-19 syndrome negatively impacts health and wellbeing despite less severe acute infection. bioRxiv. 2020; published online Nov 6. DOI:10.1101/2020.11.04.20226126.

191 Menges D, Ballouz T, Anagnostopoulos A, et al. Estimating the burden of post-COVID-19 syndrome in a population-based cohort study of SARS-CoV-2 infected individuals: Implications for healthcare service planning. 2021; published online March 1. DOI:10.1101/2021.02.27.21252572.

192 Sheehy LM. Considerations for Postacute Rehabilitation for Survivors of COVID-19. JMIR Public Health Surveill 2020; 6: e19462.

193 Haendel MA, Chute CG, Bennett TD, et al. The National COVID Cohort Collaborative (N3C): Rationale, design, infrastructure, and deployment. J Am Med Inform Assoc 2021; 28: 427–43.

